# Two modes of aversive control in suicidality: joint computational modelling exposes regime-specific clinical signatures invisible to symptom-based stratification

**DOI:** 10.64898/2026.06.09.26355278

**Authors:** Pamina Laessing, Povilas Karvelis, Ana Sophia Rashid-Cocker, Anthony C. Ruocco, Jacob W. Koudys, James Kennedy, Clement C. Zai, Peter Dayan, Andreea Diaconescu

## Abstract

Suicidal thoughts and behaviours (STBs) are heterogeneous in their proximal dynamics, planning, and stress-sensitivity, yet most subtyping efforts remain symptom-driven and rarely validated across independent datasets. Computational mixture modelling offers a principled alternative: by fitting explicit models of learning and action selection and partitioning individuals by their latent parameter profiles, it can identify mechanistically distinct control strategies invisible to cross-sectional symptom measurement. We applied this approach to aversive Go/NoGo performance, jointly clustering two independently collected STB-enriched samples (N = 50 and N = 184) using tasks with the same structure but different duration, reversal timing, and clinical instrumentation. Two recurrent behavioural regimes emerged: a fast/adaptive regime characterised by rapid policy updating and elevated feedback reactivity, and a slow/perseverative regime characterised by slow updating, high choice determinism, and a pronounced cost following contingency reversal. These regimes were stable across initialisations, recovered more parsimoniously in joint than independent solutions, and were largely orthogonal to symptom-based stratification. Critically, stratification by regime exposed clinical–computational coupling structures substantially attenuated in pooled analyses. Pooled, population-level associations were modest and anchored by a broad affective burden axis. Within the slow/perseverative regime, coupling reorganised around learning dynamics and internalizing burden (depression, hopelessness, and active suicidal ideation) with markedly larger effect sizes. Within the fast/adaptive regime, a dissociation between anxious–compulsive and antisocial–disinhibitory profiles emerged along the same computational axis, invisible at the population level. These findings support a view of suicidality heterogeneity in which clinically similar individuals differ in the control strategies they recruit under aversive uncertainty – variation that symptom measurement alone cannot capture.

## 2 Introduction

Suicidal thoughts and behaviours (STBs) arise from a complex interplay of biological, psychological, and social influences. Risk factors span demographic and sociocultural context, life stressors, and mental and physical illness, as well as cognitive traits and genetic vulnerability (e.g., (1–7). Perhaps because of this breadth, large-scale reviews link STBs to diverse neural and behavioural correlates, often implicating emotion regulation, impulse control, and decision-making systems. Yet the direction and specificity of these associations differ across disorders and study designs (3,8), and across neurobiological, neurocognitive, and clinical literature.(9,10). Reported effects vary with sampling frame (e.g., clinical vs community; ideation vs attempt), measurement window (state vs trait), and comorbidity structure. Accordingly, most established predictors are limited in their ability to forecast suicidal behaviour, particularly at the individual level (11).

A prominent response to these challenges is to look for latent substructure in suicidality, in an effort to tease apart multiple partially dissociable pathways. According to this view, STBs may differ qualitatively in various ways, including proximal dynamics (e.g., rapid fluctuations vs sustained ideation), degree of planning, and sensitivity to environmental stressors. Failing to account for this latent substructure may obscure mechanism–symptom relationships and dilutes effects (12).

Consistent with this perspective, recent work has pursued subgrouping and phenotyping using diverse approaches, including stratification based on clinical/demographic profiles, longitudinal trajectories of suicidal ideation, mechanistic proposals involving stress responsivity and impulsive aggression, and biological signatures (e.g., (6,12–18)). However, systematic, model-driven, stratification that is grounded in mechanistic, behavioural-cognitive, processes remain underdeveloped, and proposed stratifications have rarely been evaluated for their robustness across task variants and populations. Reviews of cognition of suicidal ideation (SI) and suicide attempt (SA) further note that cognitive findings often fail to generalize across psychiatric diagnoses and study contexts, limiting convergence on reliable cognitive profiles (3). Here, we address this gap by applying joint computational modelling across two independently collected datasets that share a common aversive Go/NoGo task structure but differ in task parameters, clinical instrumentation, and recruitment context – testing whether mechanistic stratification can identify robust behavioural subtypes and expose clinical–computational relationships that are undetected in pooled analyses.

Computational approaches offer a principled route to mechanistic stratification (e.g., (19–21)). By fitting explicit models of learning, action selection, and bias, they yield latent parameters that can (i) compactly summarize the generative processes underlying individual task performance, (ii) distinguish mechanistically distinct processes that produce similar observable behaviour, and (iii) suggest putative links to neurobiology and symptom dimensions. The aim is therefore not only prediction, but mechanistic interpretability, particularly when computationally defined strategies replicate across samples and relate to clinically meaningful dynamics. For example, computational accounts have proposed simulation-based subtypes of suicidality with hypothesized neurobiological correlates (22). Note that we reserve the terms ‘subtype’ for putative latent classes of suicidality that were proposed in prior work, ‘cluster’ to denote the empirical, parameter-based groupings identified in our data, and ‘regime’ to refer to the behavioural strategy profile characterising each cluster. We ultimately examine how our empirically derived clusters map onto the previously proposed subtypes.

Aversive reinforcement learning under action–valence conflict provides a particularly relevant substrate for such analyses. Go/NoGo tasks that orthogonalize action (Go vs NoGo) and valence (reward acquisition vs. punishment avoidance) offer a well-validated assay of reflexive biases and action learning (23–27). When outcomes are volatile (e.g., including a contingency reversal), these paradigms engage aversive control under changing environments, and may be experienced as stress-inducing, thereby increasing their validity as measurement instruments.

Here, we test whether clustering based on computational modelling of performance in aversive Go/NoGo tasks yields robust clusters in suicidality-relevant samples and provides a mechanistic complement to existing subtype frameworks. We address three questions:

> *(1) Can we identify clusters of strategies using computational parameters, and are these clusters sufficiently consistent across datasets to support joint modelling? Specifically, we hypothesise that, consistent with mechanistic predictions (e.g.,*(22)*), the tasks will yield ≥2 stable clusters with reproducible computational–behavioural signatures (regimes), one regime exhibiting high choice determinism and slow updating* (slow/perseverative), and one exhibiting high feedback reactivity and faster learning (fast/adaptive).
>
> *(2) Do computationally identified regimes correspond to patterns recoverable from conventional symptom-severity-based stratification, or do they capture behavioural variation that is orthogonal to clinical severity while potentially mapping onto broader phenotype profiles, as defined by characteristic behavioural, cognitive, and clinical signatures, proposed in prior suicidality research? We hypothesise that computational and symptom-severity-based clustering will be largely dissociated, while the between-regime (inter-cluster) clinical patterns will qualitatively align with previously proposed endophenotypic distinctions*.
>
> *(3) Does stratification by regime reveal within-cluster clinical-computational coupling structures that are attenuated or absent in pooled analyses? We hypothesise that regime-specific analyses will yield stronger and more interpretable associations between computational parameters and clinical variables than pooled analyses, consistent with mixture cancellation masking regime-specific relationships in unsegregated samples*.

To address these questions, we apply model-dependent stratification of aversive Go/NoGo performance using empirical hierarchical Bayesian inference (eHBI;(28,29)) mixture modelling on two independently collected datasets that share a common task structure but differ in task duration, reversal timing, and clinical instrumentation. Critically, we performed the clustering jointly across both datasets, enforcing a shared computational structure, while allowing dataset-specific parameter distributions. This joint approach provides a direct test of whether the identified regimes reflect a common computational phenotype rather than dataset-specific artefacts. To characterise the relationship between these regimes and clinical variables, we further apply a joint canonical correlation analysis (CCA) that constrains the computational weight vector to be shared across datasets while permitting dataset-specific clinical representations. Across both datasets, the joint modelling identifies two recurring strategy regimes with reproducible computational–behavioural signatures. This stratification has potential to expose regime-dependent clinical–computational coupling, distinct patterns of association between clinical dimensions and mechanistic parameters, that is attenuated or obscured in pooled analyses.

## 3 Methods

### 3.1 Datasets and Task Designs

We analysed two independently collected datasets comprising clinical and help-seeking community-recruited samples with varying histories of suicidal thoughts and behaviours (STBs), including suicidal ideation (SI) and suicide attempt (SA). The datasets shared the administration of an aversive Go/NoGo task with a state-action reversal but differed in some of the task implementation details and clinical questionnaires that were administered to participants. An overview of samples and task variants is provided in Figure 2-1.

**Figure 3-1.**
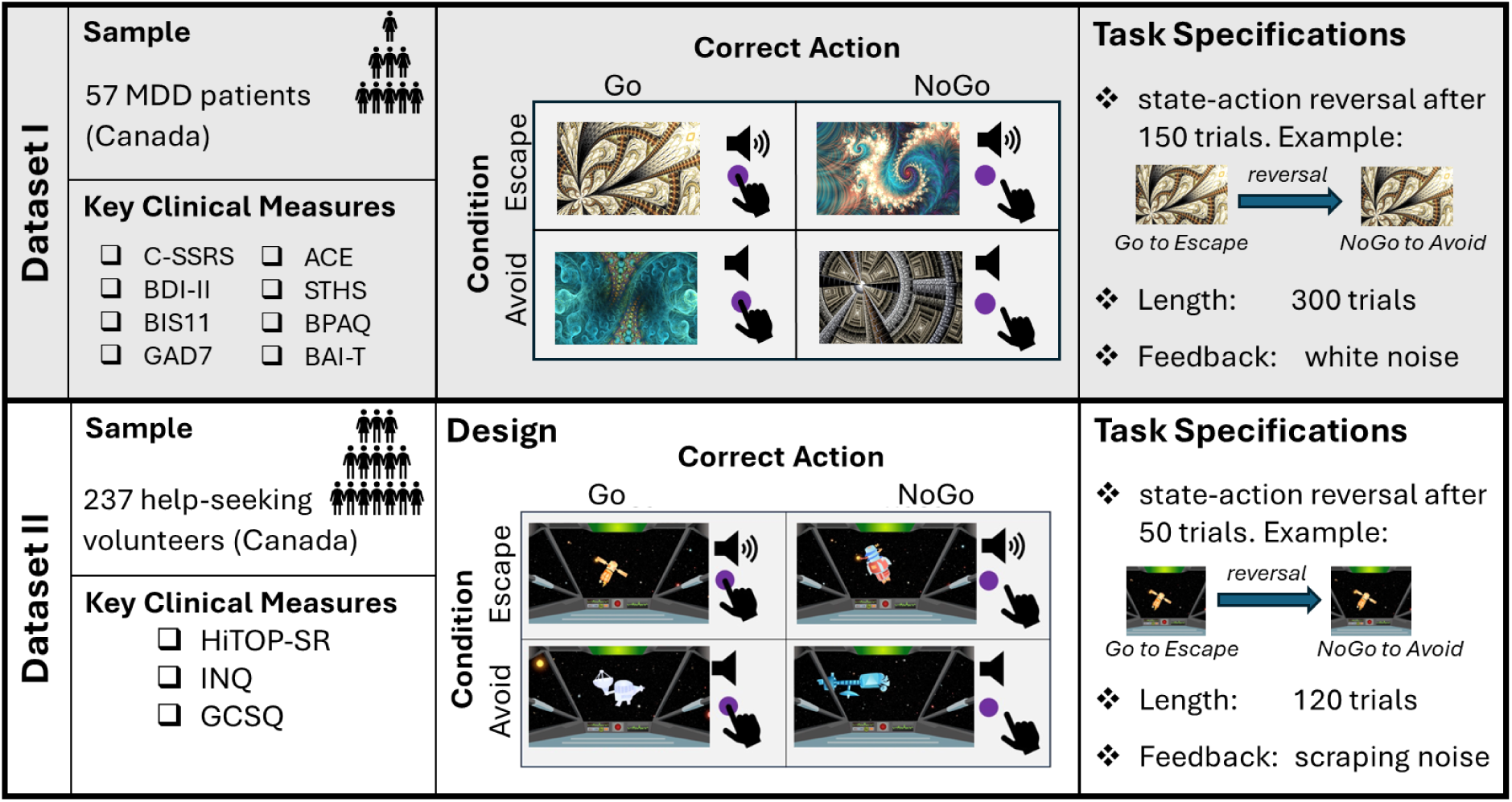
Overview of datasets and task designs for the aversive Go-NoGo task. *Abbreviations: MDD: major depressive disorder, C-SSRS: Columbia Suicide Severity Rating Scale, BDI-II: Becks Depression Inventory, BIS11-Barrat Impulsiveness Scale, GAD7: Generalizied Anxiety Disorder Scale-7, ACE: Adverse Childhood Experience, STHS: State-Trait Hopelessness Scale, BPAQ: Buss Perry Aggression Questionnaire, BAI-T: Beck’s Anxiety Inventory-Trait, HiTOP-SR: Hierarchical Taxonomy of Psychopathology, Self-Report, INQ: Interpersonal Needs Questionnaire, GCSQ: German Capability for Suicide

#### 3.1.1 Populations

##### Dataset I

Participants were drawn from a clinical study aimed at identifying suicidal subgroups based on behavioural, psychological, and genomic factors. We analysed an early subsample of this study. Participants were re-contacted from IMPACT (Individualized Medicine: Pharmacogenetic Assessment and Clinical Treatment), an ongoing provincially funded pharmacogenetics study (30). The IMPACT cohort included adults with diagnoses including anxiety, depression, psychosis, pain, ADHD, bipolar disorder, tic disorders, and sleep disorders.

Participants were eligible for the suicide-focused study if they were at least 18 years of age and had a clinical diagnosis of mood disorder. Ineligible participants were under 18 years of age, currently hospitalized, had psychosis or schizoaffective disorder diagnoses, were unable to provide informed consent. Participants completed study assessments, including the Go/NoGo task. Of 57 participants who completed the task, 50 (72% female, mean age: 39.6±12.4) were retained based on outlier performance measures. (see Supporting Information 1.1.1)

Suicidality was assessed using the Columbia-Suicide Severity Rating Scale (C-SSRS; (31)). Additional self-report measures indexed depressive symptoms (BDI-II: Beck’s Depression Inventory;(32,33)), hopelessness (STHS: State-Trait Hopelessness Scale; (34)), anxiety symptoms (GAD-7: Generalizied Anxiety Disorder Scale-7;(35) ; BAI-T: Beck Anxiety Inventory – Trait (36)), impulsivity (BIS11: Barrat Impulsiveness Scale; (37))), aggression (BPAQ: Buss Perry Aggression Questionnaire; (38)) and childhood trauma (ACE: Adverse Childhood Experience; (39)). Of the analyzed participants, 27 reported SI, and 23 reported SI and SA within their lifetime, with 3 participants reporting recent SA (past year). The online Go/NoGo task employed included 300 trials, with choice and reaction time recorded.

##### Dataset II

Dataset II comprised adults recruited via Prolific in Canada and the US who self-reported a current or past mental health diagnosis on the platform. Participants completed a battery of five cognitive tasks including the Space Explorer Go/NoGo task and were assessed for suicidality with the German Clinical Suicidality Questionnaire (GCSQ, (40)) and Interpersonal Needs Questionnaire (INQ, (41)). Additionally, all participants completed the self-report questionnaire for the Hierarchical Taxonomy of Psychopathology (HiTOP-SR, (42)). Of 254 recruited participants, 236 completed the Go/NoGo task; after exclusion, 184 participants were included in the current analyses (75% female, mean age: 34.6±10.3).

##### Ethics Statement

The IMPACT study (Dataset I) was approved by the Centre for Addiction and Mental Health Research Ethics Board (REB Number: 021/2016). All participants provided written informed consent to participate in this study. Data analysed in this paper was collected between December 2, 2021, and May 17, 2023.

Dataset II was conducted online via Prolific and was approved by the University of Toronto Social Sciences, Humanities and Education Research Ethics Board (RIS Protocol Number: 40050). All participants provided informed written consent electronically prior to participation. Data analysed in this paper was collected between September 14, 2024, and October 8, 2024.

#### 3.1.2 Cognitive task design

We used orthogonalized aversive Go/NoGo tasks to quantify reflexive action–valence tendencies in the context of suicidality. Orthogonalizing action (Go vs NoGo) and valence framing (escape vs avoid) enables estimation of action–valence coupling (“Pavlovian bias”), expressed as facilitated performance in Pavlovian-congruent conditions and impaired performance in incongruent conditions (23,43–45).

Across task variants, participants learned to select the optimal response (Go vs NoGo) for each cue to minimize aversive outcomes by trial and error. Four cues each corresponded to one condition: Go-to-Escape, Go-to-Avoid, NoGo-to-Escape, and NoGo-to-Avoid. In **escape** trials, an aversive sound was played during cue presentation and could be terminated by the correct response. In **avoid** trials, no sound was played during cue presentation; depending on the action, an aversive sound could be triggered. Outcome contingencies were probabilistic, with optimal responses resulting in silence 80% of the time, the remaining 20% resulting in the aversive stimulus. Suboptimal responses reversed these probabilities.

Trial structure parameters (cue duration, response window, feedback duration, and inter-trial interval) differed across datasets and are reported in Supporting Information table 1. Behavioural outcomes included trial-wise choices and reaction times.

**Table 1.**
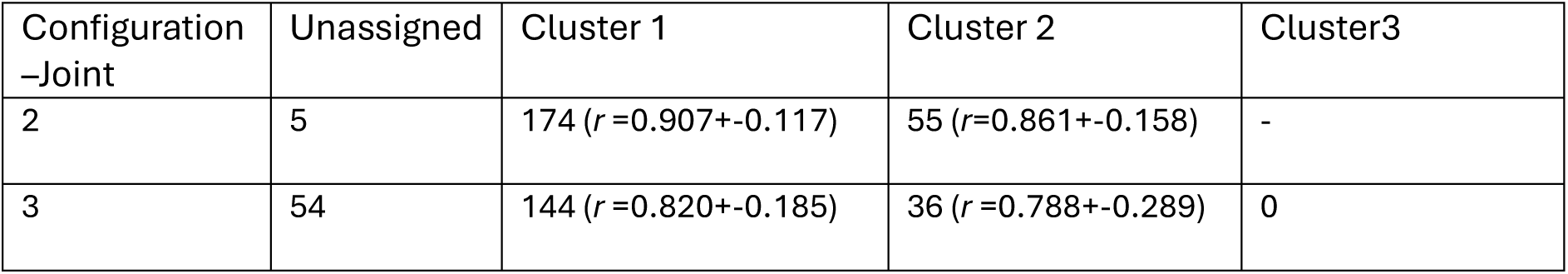
Cluster assignments for joint datasets I and II with 2 and 3 clusters respectively. The numbers of individuals unassigned or reliably assigned to one of the clusters as per majority voting (>=6/8 runs) are shown. The assignment responsibilities r of individuals to cluster are averaged across 8 runs (mean and standard deviation).

Two task implementations were used, differing in total number of trials, feedback stimuli, and reversal structure.

##### Dataset I (300 trials; reversal)

The task comprised 300 trials, with a reversal after 150 trials in which cues were reassigned to conditions (i.e., the cue–action and cue–valence mapping inverted; for example, a cue previously requiring Go-to-Avoid required NoGo-to-Escape after reversal). Conditions were cued by four visually distinct fractals. Aversive feedback consisted of white noise; participants were instructed to adjust headphone volume to an unpleasant but tolerable level. Subjective probes (e.g., perceived difficulty and controllability) were included. Trials were pseudo-randomized to be approximately balanced across cues within each block (i.e., NoGo-to-Avoid = 67, Go-to-Avoid = 79, NoGo-to-Escape = 78, Go-to-Escape = 76).

##### Dataset II (120 trials; reversal; gamified)

The task comprised 120 trials and included a reversal after 50 trials in which cues were reassigned to conditions (as in the variant for Dataset I). The task was presented as a gamified “Space Explorer” paradigm: on each trial, participants decided whether to shoot approaching space debris (Go) or let it pass (NoGo). In escape conditions, the approaching space debris was accompanied by the unpleasant sound, while avoid conditions were silent. Aversive feedback consisted of a screeching sound and an explosion animation; neutral feedback was silence and explosion debris disappeared in smoke. Participants were instructed to adjust headphone volume to an unpleasant but tolerable level. A pre-generated randomized sequence ensured equal cue counts (i.e., 30 each) across 120 trials.

### 3.2 Modelling and fitting

#### 3.2.1 Behavioural Data Analyses

Behavioural data were initially analysed using both task-derived metrics and simple model-dependent heuristics (Win-Stay and Lose-Switch strategies). We computed accuracy (the ratio of correct and incorrect responses across all trials) on the overall task, as well as for each of the four cue types, and before and after the reversal for Dataset I and II. A behavioural Go bias was defined as the difference in accuracy for Go trials (Go-to-Avoid and Go-to-Escape) relative to NoGo trials (NoGo-to-Avoid and NoGo-to-Escape). In line with the orthogonalized Go/NoGo literature (e.g. (23–25,43)), we operationalized Pavlovian performance biases as condition-specific advantages for Go/NoGo choices. These measures reflect the performance difference between Pavlovian-congruent and Pavlovian-incongruent mappings within each valence context. Specifically, the Go-to-Escape bias was calculated as the accuracy difference between Go and NoGo trials in the Escape condition. The NoGo-to-Avoid bias was calculated as the accuracy difference between NoGo and Go trials in the Avoid condition. Positive values of the Pavlovian performance bias therefore indicate a performance advantage for Pavlovian-congruent action mappings within the corresponding context.

#### 3.2.2 Computational model

We created a candidate model set (12 models, varying mechanisms for interactions among learning, reward sensitivity, forgetting, and Pavlovian bias; including 11 RL models and one based on active inference). We adapted the popular empirical HBI (eHBI) procedure to improve its convergence on the present data and model space (28) and applied it separately to each dataset to select the best-fitting model based on model evidence and protected exceedance probability. EHBI allows us to compare candidate models while accounting for inter-individual variability (29).

Across both datasets, one particular model (RL-Pf) clearly fit the best, with no indication that a competing model better accounted for a meaningful subset of participants. Its exceedance probabilities were *p*_*e*,*I*_ = .98, *p*_*e*,*II*_ = 1 (see Supporting Information 1.2).

The RL-Pf model has three components: instrumental state-action values *Q*(*s*, *a*) that are updated according to a Rescorla-Wagner rule but are subject to forgetting and decay; Pavlovian bias terms that are fixed across trials; and a softmax-based action selection mechanism with no lapse.

##### State-action value learning

The RL-Pf model uses a simple delta-rule (Rescorla–Wagner) to update state–action values *Q*(*s*, *a*). After taking action *a*_*t*_ in state *s*_*t*_ and observing outcome *r*_*t*_, the chosen action value was updated as:

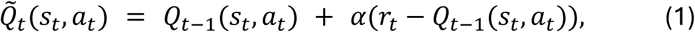

where *α* ∈ [0,1] is the learning rate. Outcomes were coded as *r*_*t*_ = 0 for a neutral outcome and *r*_*t*_ = −1 for an aversive outcome. All other state-action values were left unchanged *Q̃*_*t*_(*s*, *a*) = *Q*_*t*−1_(*s*, *a*), *where s* ≠ *s*_*t*_, *a* ≠ *a*_*t*_.

##### Forgetting/decay mechanism

All the state-action values (for both hosen and unchosen actions) undergo multiplicative decay (with parameter *ϕ* ∈ [0,1]) after each trial, capturing forgetting/interference over the course of the task (46):

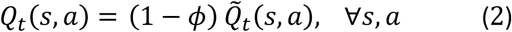

##### Pavlovian bias terms

Pavlovian tendencies were modeled as fixed additive biases on top of the state–action values (26,47). Biases were added to learned values to form final action propensities *W*:

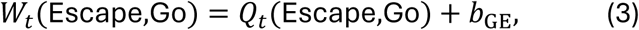

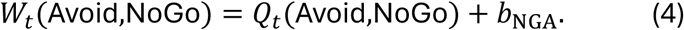

As instrumental learning progresses, the relative contribution of fixed bias terms to choice depends on the magnitude of learned values *Q*.

##### Action selection

To generate choice probabilities, we used a SoftMax rule with inverse temperature *β* applied to final action propensities *W*_*t*_(*s*, *a*):

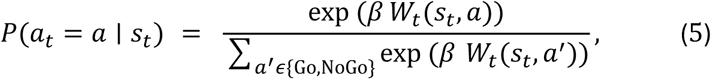

where larger *β* yields more deterministic choices.

##### Model-driven subgrouping with empirical hierarchical Bayesian inference

Having used eHBI to perform model comparison across all participants in each dataset to determine the best model, we then used it again for the further analyses.

###### Stage 1: Subgrouping as mixtures of the winning model

To test whether each population could be partitioned into subgroups under a shared computational mechanism, we re-ran eHBI across both datasets. Here, we fit mixtures with *K* ∈ {1,2,3} latent clusters, where each cluster was modeled using RL-Pf but allowing cluster-specific (but dataset general) parameter priors (“*K* instances” of RL-Pf). This approach targets clusters that share the same model structure (RL-Pf) but differ in parameter regimes. We limited *K* ≤ 3 given the sizes of our datasets.

###### Stage 2: Cluster number selection and stability criteria

To determine the number of latent regimes, we compared configurations using model evidence, Bayesian Omnibus Risk (BOR), and integrated Bayesian Information Criterion (iBIC; (48)), together with assignment robustness across random initializations. *Model evidence* (marginal likelihood) quantifies the integrated probability of the data under a configuration; higher values indicate better support. BOR is the posterior probability that apparent model-frequency differences could be explained by chance (i.e., that there is no real difference across candidate configurations); lower BOR therefore indicates stronger evidence that the favoured configuration is meaningfully better. iBIC provides an evidence-approximating information criterion that penalizes complexity; lower iBIC indicates a preferred trade-off between fit and parsimony.

For each configuration, we assessed robustness over 8 random initializations. Parameters were converted from Gaussian distributions across the population to [0,1] using a logistic sigmoid transformation, where we randomized the starting points of each fitted parameter value with a deliberately broad *x*_*init*_ = Gaussian(*μ* = 0, *σ*^2^ = 4). Participants were labelled by maximum responsibility (*r*; posterior membership probability) within each fit, and stability was quantified by majority voting across runs. Given that cluster labels are not preassigned (i.e. cluster 1 in run 1 may correspond with cluster 2 in run 2), we employed a maximum assignment permutation approach to align labels. The approach was verified based on group parameter assignment for each cluster. A participant was considered stably assigned if the same cluster label was obtained in ≥ 6/8 fits; otherwise, the participant was treated as lacking a stable label for that configuration. We then selected the smallest *K* that had strong Bayesian support (higher evidence / lower iBIC and low BOR for the improvement over simpler models) and demonstrated high assignment stability and interpretability. Configurations that improved Bayesian diagnostics but produced substantial instability (i.e., >20% of participants failing the stability criterion and/or systematically lower responsibilities) were treated as over-fragmented and were not retained for downstream analyses.

#### 3.2.3 Statistical analysis for cluster interpretation

Analyses were conducted to perform basic model validation checks, characterize between-cluster differences in cognitive–behavioural features, and examine exploratory symptom–behaviour coupling within clusters.

##### Model validation

Parameter recoverability was evaluated as a simulation-based sanity check: we generated data from the hierarchically fitted parameters and refit the model to the synthetic data, quantifying agreement between generating and recovered parameters using intraclass correlation coefficients (ICC) across all fitted parameters. Parameter interpretability (i.e., a posterior predictive check) was assessed using Pearson correlations between task-derived behavioural measures and fitted computational parameters.

##### Between-cluster comparisons

To characterize the cognitive–behavioural features underlying subgrouping, we tested between-cluster differences in task-derived behavioural measures (overall Go bias, NoGo-to-Avoid performance, Go-to-Escape performance, lose–switch, win–stay) using two-sided independent-samples t-tests. We additionally used two-sided t-tests to assess between-cluster differences in clinical and demographic variables to identify obvious stratification factors.

##### Within-cluster symptom–behaviour coupling

To examine symptom–behaviour coupling within and across clusters, we implemented a joint canonical correlation analysis (CCA) that enforces a shared computational weight vector across datasets while permitting dataset-specific clinical representations. This approach was chosen because direct one-to-one mappings between individual symptoms and computational parameters could not be assumed *a priori*; and because the clinical instruments differed across datasets, precluding direct variable harmonisation. The joint formulation instead identifies the shared computational axis or axes that best explain clinical variance in both datasets simultaneously, while allowing the clinical projection to differ.

##### Clinical dimensionality reduction

Prior to CCA, clinical variables were subject to dimension reduction separately within each dataset using exploratory factor analysis (EFA). Variables were standardised within dataset before EFA. Factors were extracted using maximum likelihood estimation; the number of factors was determined by parallel analysis; and an oblique rotation was applied to allow for correlated factors. The resulting factor scores constituted the dataset-specific clinical matrices ***Y***_***d***_ entering the joint CCA. Available demographic variables were included with the clinical variables: Dataset I (age, self-identified sex) and Dataset II (age, sex assigned at birth, medication status). Computational parameters were standardised within each dataset before CCA.

##### Joint CCA formulation

Let **X**_*d*_ ∈ ℝ^*n_d_*×*p*^ denote the matrix of the *p* computational parameters, and *q*_*d*_ ∈ ℝ^*n*_*d*_×*q*_*d*_^ denote the matrix of the *q*_*d*_ EFA factor scores for the *n*_*d*_ members of dataset *d* ∈ {*I*, *II*}. The computational parameter space is identical across datasets (*p* shared), whereas the clinical feature space may differ (*q*_1_ ≠ *q*_2_). All variables were z-scored within dataset prior to analysis. For each canonical component *j*, we estimate a shared parameter-side weight vector ***a***^(*j*)^ ∈ ℝ^*p*^, and dataset-specific clinical weight vectors 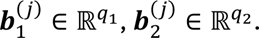 The corresponding canonical variates are:

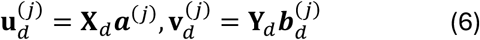

Rather than maximising Pearson correlations, the joint model is formulated in terms of cross-covariance bilinear forms. Specifically, for each component, we solve:

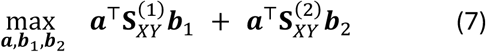

where 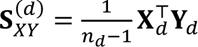 denotes the empirical cross-covariance matrix in dataset *d*. Both datasets contribute equally to the objective. The optimisation is subject to variance normalisation constraints:

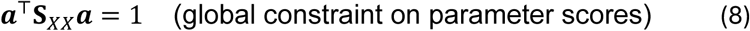

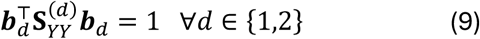

, where 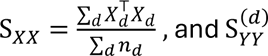 are the within-dataset covariance matrices. The constraint on ***a*** enforces a shared, scale-normalised computational axis, while clinical variates are normalised separately within each dataset.

The optimisation was performed using an iterative alternating scheme, updating *a*, *b*_1_, and *b*_2_in turn until convergence. Following estimation of each component, deflation was performed to remove the variance explained by the extracted canonical variates from both X_*d*_ and Y_*d*_, and the procedure was repeated to obtain subsequent orthogonal components.

The joint CCA was applied at two levels. At the population level, the model was fit across all participants in both datasets to identify global computational–clinical dimensions. To characterise heterogeneity identified through computational clustering, the same procedure was applied within each cluster separately, yielding cluster-specific canonical dimensions and enabling assessment of whether distinct computational–clinical coupling structures emerge across regimes.

##### Relationship to existing multi-set association methods

The joint CCA used here is related to generalised and linked CCA approaches (49,50) but differs in enforcing an asymmetric constraint: the computational weight vectors are shared across datasets while clinical weight vectors are free to differ. This asymmetry reflects a specific scientific assumption, that the computational parameters index the same latent constructs across datasets while the clinical instruments need not (in our case, since different clinical measurements were taken). Multi-block p variants (e.g., JIVE, OnPLS; (51,52)) offer an alternative decomposition framework but do not naturally accommodate one-sided sharing constraints of this kind.

##### Bootstrapping for robustness

To assess the stability of the exploratory factor analysis–canonical correlation analysis (EFA–CCA) results, we implemented a non-parametric bootstrap procedure, an approach with an established history in the CCA literature (53–55). The bootstrap was performed separately for each CCA analysis (population-level and cluster-specific analyses). In each bootstrap iteration, participants were resampled with replacement from the original dataset while maintaining the original sample size. The full analysis pipeline was then re-estimated on the resampled dataset, including (i) exploratory factor analysis of the clinical variables and (ii) canonical correlation analysis between the resulting clinical factors and the computational parameters. For each bootstrap iteration, we recorded canonical correlations, variance explained by each canonical variate, associated significance tests, and the correlations (loadings) between variables and canonical variates. This procedure was repeated for *N* = 1000 bootstrap iterations. For each bootstrap sample, variables were ranked according to the magnitude of their association with the canonical variates (i.e., their loadings/correlations with the canonical dimension). Rankings were computed separately for computational parameters and clinical variables and for each canonical variate. Across bootstrap iterations, two summary metrics were calculated for each variable: (i) the mean rank, representing the average position of the variable across bootstrap samples (with rank 1 indicating the strongest association with the canonical variate), and (ii) the percentage of iterations in which the variable was ranked first/top 10% /top 20%, reflecting how frequently it emerged as the dominant contributor. These summaries were used to characterize the stability and relative prominence of variables contributing to each canonical variate.

#### 3.2.4 Clinical-variable clustering analysis

To examine whether the behavioural phenotypes identified through computational modelling correspond to patterns captured by conventional symptom-based stratification, we performed an additional clustering analysis based solely on clinical variables.

For each dataset, all available clinical measures (including HiTOP subfactors where available, and suicidality-related variables such as INQ and GCSQ measures) were used as input features. Prior to clustering, variables were standardized (z-scored) within each dataset to ensure comparability across scales.

Participants were clustered using k-means clustering applied to the standardized clinical variables. The optimal number of clusters was determined separately for each dataset using the elbow method based on the within-cluster sum of squares.

After identifying clinically derived clusters, the reinforcement-learning model used in the primary analyses (RL-Pf) was hierarchically fit separately within each clinical cluster to estimate computational parameters governing learning dynamics, Pavlovian biases, and action selection. Parameter estimates between clusters were compared using two-sample *t*-tests, with Bonferroni correction applied across parameters.

To assess the relationship between symptom-based and computational stratification, we examined the distribution of computational cluster assignments within the clinically derived clusters. This allowed us to evaluate whether symptom-based clustering reproduced the computationally identified behavioural regimes or instead captured an orthogonal grouping structure.

## 4 Results

We report findings in four steps. First, we characterise task performance and baseline action–valence asymmetries across both datasets. Second, we identify the computational regimes, evaluating cluster selection, assignment stability, parameter recoverability, and parameter interpretability, and confirm that a shared two-regime structure fits both datasets more parsimoniously than independent solutions. Third, we characterise the identified regimes in terms of their computational and behavioural signatures and directly test whether these regimes correspond to patterns recoverable from symptom-based stratification alone. Fourth, we examine clinical–computational coupling at the population level and within each regime, testing whether stratification exposes associations that are attenuated in pooled analyses. Cross-dataset correspondence is evaluated throughout to assess whether the identified regimes reflect a shared computational structure rather than dataset-specific patterns.

### 4.1 Task Performance and Behavioural Signatures

**Figure 4-1.**
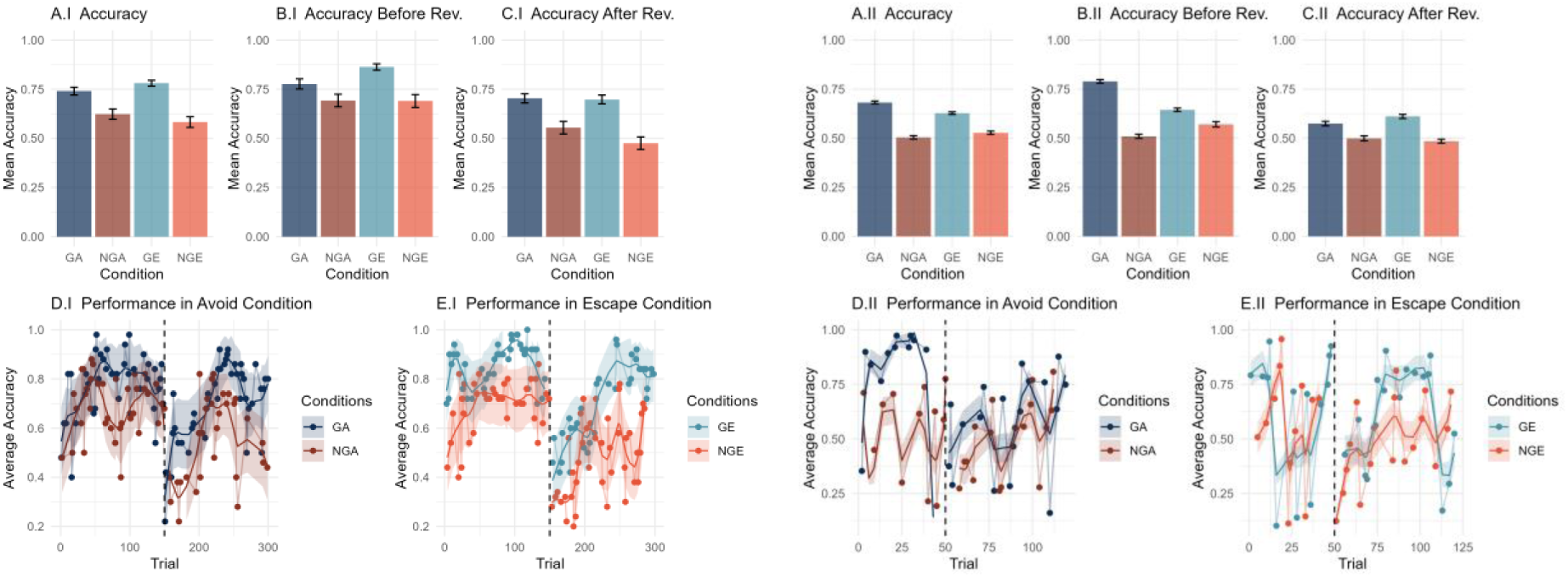
Accuracy of performance on the tasks. The left panels (A.I-E.I) correspond to dataset I, the right panels (A.II-C.II) correspond to dataset II. (A-C) Overall accuracy and accuracy before and after reversal, split by condition. Task performance is dominated by a Go-bias, where accuracy was higher for Go than NoGo cues during both conditions. (D-E) average accuracy per trial (dots; the lines show the result of spline smoothing, separately applied before and after the reversal; dashed line at 150 (50) indicates the reversal). Accuracy initially increases across conditions, and we observe a clear decline immediately after the cue-outcome reversal, followed by noisy learning. Error bars represent standard error of the mean and shaded areas indicate a 95% confidence interval. GA = Go-to-Avoid; NGA = NoGo-to-Avoid; GE = Go-to-Escape; NGE = NoGo-to-Escape.

#### Dataset I

Participants achieved an average accuracy of 68.2% (SD = 10.7). A repeated-measures ANOVA on accuracy with factors condition (Escape vs Avoid), response type (Go vs NoGo), and phase (pre- vs post-reversal) indicated main effects of response type and phase (p = 9.7×10e^−15^ and p = 2.7×10e^−13^, respectively), with higher accuracy for Go responses and prior to reversal. A response type × condition interaction was also observed (p = .039), indicating context-dependent modulation of Go/NoGo performance.

Bias indices showed *b*_*GE*_ = 0.206 (SD = 0.205) and *b*_*NGA*_ = −0.107 (SD = 0.221). Under this definition, the negative *b*_*NGA*_ indicates lower accuracy in NoGo-to-Avoid relative to Go-to-Avoid at the population level. However, relative to the Avoid condition, the Go bias in the Escape condition was significantly higher (p = .004), consistent with an inhibitory-avoid bias despite the negative *b*_*NGA*_ value.

#### Dataset II

Following exclusion criteria, the average accuracy was only 57.8% (SD = 5.87). A repeated-measures ANOVA indicated a small main effect of condition (p = .049), and significant effects involving phase (pre-/post-reversal) and response type (p < .001). Consistent with this, participants showed higher accuracy for Go than NoGo trials. Bias indices were *b*_*GE*_ = 0.107 (SD = 0.148) and *b*_*NGA*_ = −0.156 (SD = 0.163), again suggesting a pronounced overall Go advantage independent of condition. No inhibitory-avoid bias was observed in this dataset.

To facilitate cross-dataset comparison, we summarize (i) overall accuracy, (ii) pre/post reversal performance, and (iii) task-derived Go bias, Pavlovian congruency indices, and lose–switch/win–stay (Figure 3-2, Supporting Information table 2). We find that the behaviours only differ in accuracy in Escape Bias (total accuracy (*p*_*bonf*_ = 3.60*e*^−8^, *d* = 0.943), accuracy-before (*p*_*bonf*_ = 2.07*e*^−8^, *d* = 0.959) and Pav-GE (*p*_*bonf*_ = 3.38*e*^−2^, *d* = 0.449)

**Figure 4-2.**
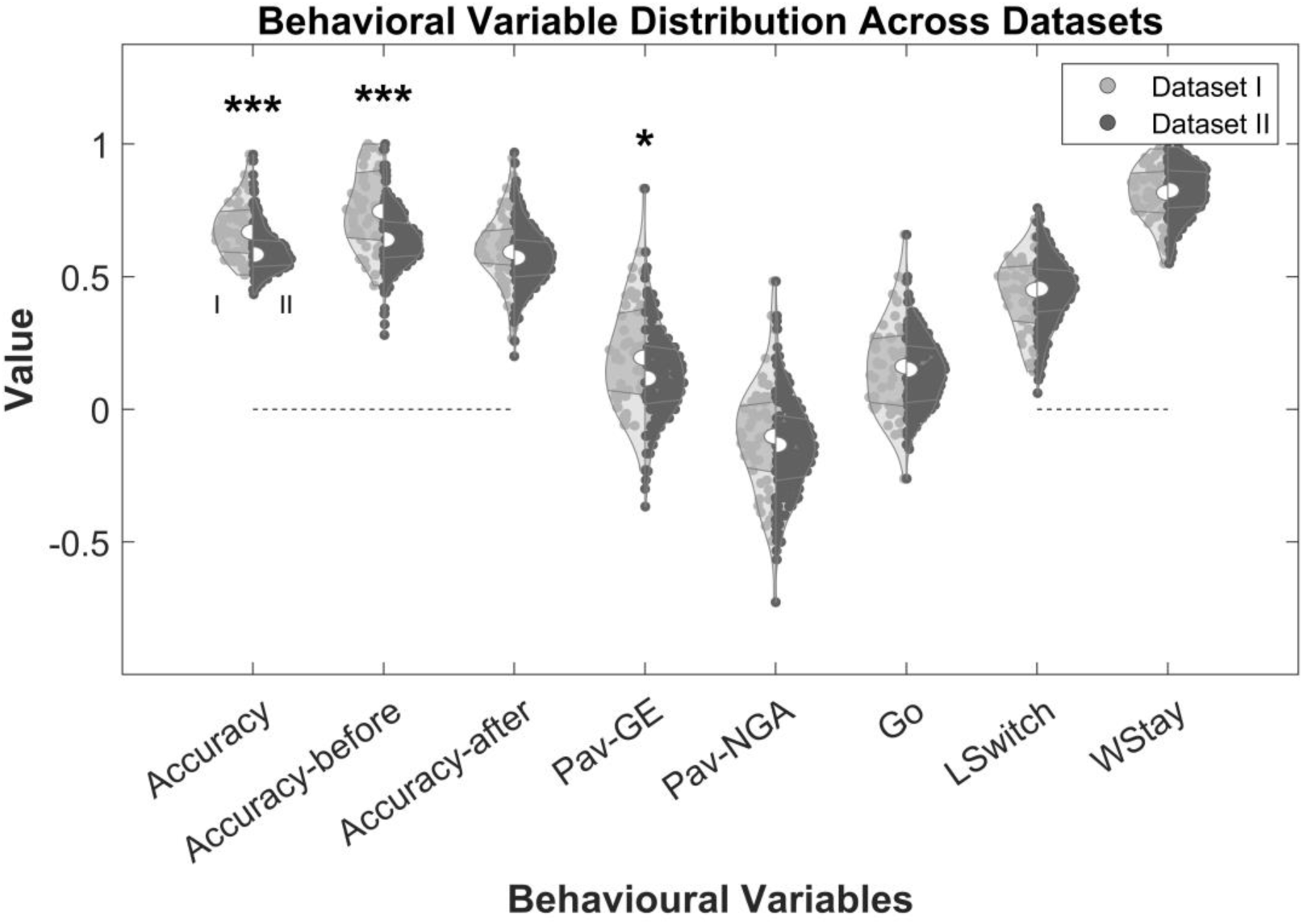
Task-derived behavioural measures compared across datasets, including performance biases (overall Go-Bias, Excitatory-Escape Bias and Inhibitory-Avoid Bias), learning strategies (Lose-Switch and Win-Stay behaviours) and accuracy (total, before and after reversal). Violins are split to compare measures across datasets with dataset I represented in the left violin halves and dataset II represented in the right violin halves. Median, 25^th^ and 75^th^ percentile is indicated for each violin half. Dotted lines at x=0 indicate measures that cannot be smaller than zero. Differences across datasets are tested with two-sided t-tests and evaluated after Bonferroni correction. GE = Go-to-Escape; NGA = NoGo-to-Avoid; LSwitch = Lose–Switch; WStay = Win–Stay, * indicates p<0.05, ** indicates p<.01, *** indicates p<.001.

**Table 2.** Cluster assignments for Dataset I with 2 and 3 clusters. The numbers of individuals unassigned or reliably assigned to one of the clusters as per majority voting (>=6/8 runs) are shown. The assignment responsibilities r of individuals to cluster are averaged across 8 runs (mean and standard deviation).

| Configuration – Dataset I | Unassigned | Cluster 1 | Cluster 2 | Cluster3 |
| --- | --- | --- | --- | --- |
| 2 | 0 | 29 ($r = 0.961\pm0.077$) | 21 ($r=0.916\pm0.120$) | - |
| 3 | 12 | 11 ($r = 0.774\pm0.317$) | 19 ($r = 0.740\pm0.383$) | 8 ($r = 0.711\pm0.409$) |

### 4.2 Cluster Selection Across Datasets with eHBI: Computational Regime Identification

We first report the joint clustering solution across both datasets, then the single-dataset solutions, and finally compare them. The joint solution is treated as primary throughout.

Given the common task structure, we examined whether the datasets can reasonably be modelled together using the overall best-fitting model RL-Pf (see Methods). To this end, we first assessed the optimal number of clusters for the joint modelling as well as the separate dataset fits. We selected the number of regimes (*K*) using the pre-specified criteria described in Methods (Section 2.2.3), combining model evidence, BOR, iBIC, and assignment stability across random initializations.

We then performed model comparison between the joint and single modelling to understand whether casting the datasets into common clusters has a reasonably good fit compared to separately defined clusters per dataset.

**Joint Modelling,** Bayesian diagnostics favored *K* > 1 configurations (config 2 vs 1: BOR = 1.9125e^−28^; config 3 vs 2: BOR = 0.0183). *K* = 2 yielded the most stable solution, with high assignment probabilities and only five participants not robustly assigned to a cluster. In contrast, the *K* = 3 configuration produced two clusters that largely replicated the *K* = 2 solution but resulted in a substantially larger number of participants without clear cluster assignment, indicating reduced stability. Cluster sizes and assignment probabilities for the joint fits are summarized in Table 3. We therefore retained the two-cluster solution for subsequent comparisons.

**Table 3.** Cluster assignments for Dataset II with 2 and 3 clusters respectively. The numbers of individuals unassigned or reliably assigned to one of the clusters as per majority voting (>=6/8 runs) are shown. The assignment responsibilities r of individuals to cluster are averaged across 8 runs (mean and standard deviation).

| Configuration – Dataset II | Unassigned | Cluster 1 | Cluster 2 | Cluster3 |
| --- | --- | --- | --- | --- |
| 2 | 9 | 121 ($r = 0.840 \pm 0.225$) | 54 ($r = 0.889 \pm 0.159$) | - |
| 3 | 42 | 112 ($r = 0.848 \pm 0.187$) | 30 ($r = 0.819 \pm 0.189$) | 0 |

#### 4.2.1 Single dataset fits

In *Dataset I*, Bayesian diagnostics favored higher-*K* configurations (config 2 vs 1: BOR = 1.7573*e*^−12^; config 3 vs 2: BOR = 3.6571*e*^−15^). However, the *K* = 3 solution was substantially less robust across runs: 24% of participants lacked a stable label under the ≥6/8 criterion, and mean posterior responsibilities were lower than in the *K* = 2 solution. We therefore retained *K* = 2 for subsequent analyses (Table 4).

**Table 4.**
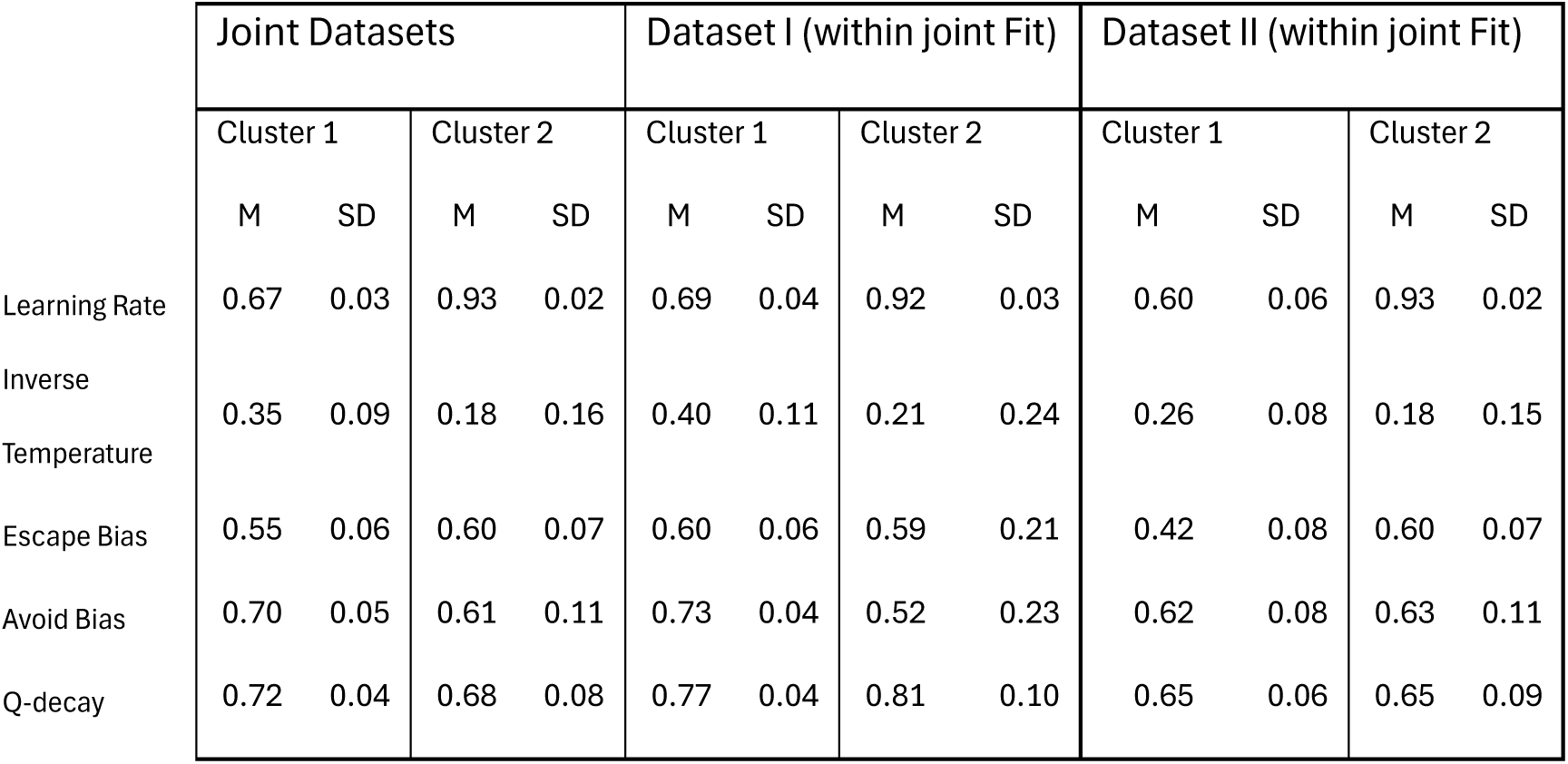
ICC. Parameter recoverability for each identified cluster. Synthetic data were generated from the hierarchically fitted group posteriors (single simulation run) and refitted with the same model. ICC values are reported for each cluster both, for joint datasets and separately for each dataset. M: mean, SD: standard deviation

|  | Joint Datasets |  |  |  | Dataset I (within joint Fit) |  |  |  | Dataset II (within joint Fit) |  |  |  |
| --- | --- | --- | --- | --- | --- | --- | --- | --- | --- | --- | --- | --- |
|  | Cluster 1 |  | Cluster 2 |  | Cluster 1 |  | Cluster 2 |  | Cluster 1 |  | Cluster 2 |  |
|  | M | SD | M | SD | M | SD | M | SD | M | SD | M | SD |
| Learning Rate | 0.67 | 0.03 | 0.93 | 0.02 | 0.69 | 0.04 | 0.92 | 0.03 | 0.60 | 0.06 | 0.93 | 0.02 |
| Inverse Temperature | 0.35 | 0.09 | 0.18 | 0.16 | 0.40 | 0.11 | 0.21 | 0.24 | 0.26 | 0.08 | 0.18 | 0.15 |
| Escape Bias | 0.55 | 0.06 | 0.60 | 0.07 | 0.60 | 0.06 | 0.59 | 0.21 | 0.42 | 0.08 | 0.60 | 0.07 |
| Avoid Bias | 0.70 | 0.05 | 0.61 | 0.11 | 0.73 | 0.04 | 0.52 | 0.23 | 0.62 | 0.08 | 0.63 | 0.11 |
| Q-decay | 0.72 | 0.04 | 0.68 | 0.08 | 0.77 | 0.04 | 0.81 | 0.10 | 0.65 | 0.06 | 0.65 | 0.09 |

In *Dataset II*, model evidence favored larger *K* values (config 2 vs 1: BOR = 3.2394*e*^−23^; config 3 vs 2: BOR = 3.3130*e*^−25^), but, again, the additional cluster in *K* = 3 did not lead to the recovery of an additional stable cluster (Table 3). Instead, the recovered clusters are largely consistent with the *K*=2 solution (110/112 were cluster 1 in *K*=2 and 30/30 were cluster 2 in *K*=2) and a total of 42 participants could not robustly be assigned to a cluster across runs (22.5% of the population). Given this pattern, we retain the *K* = 2 solution for analysis.

#### 4.2.2 Model Comparison – Joint vs separate modelling

To evaluate whether a shared cluster structure provided a more parsimonious account of the data than independently derived solutions, we compared the aggregate model fit of the joint clustering approach to that of separate per-dataset fits. For each solution, we computed an integrated Bayesian Information Criterion (iBIC) score aggregated across both datasets, alongside dataset-specific approximations.

The joint two-cluster solution yielded a marginally lower aggregate iBIC than the joint score of the two independent fits (joint: 7.1808e⁴, 306.87 per subject; separate: 7. 1874e⁴, 307.15 per subject). At the dataset level, per-dataset iBIC scores were slightly lower for the independent fits (Dataset I: 2.8658e⁴ vs. 2.8780e⁴; Dataset II: 4.3634e⁴ vs. 4.3699e⁴ for separate vs. joint, respectively). This pattern at dataset level is expected: fits optimised within a single dataset will always better capture local idiosyncrasies. Critically, however, the aggregate advantage of the joint solution indicates that a shared two-cluster structure accounts for the joint data more parsimoniously, without meaningful information loss at the level of individual datasets. We therefore treat the joint clustering solution as the primary analysis for all subsequent comparisons.

#### 4.2.3 Parameter validation (recoverability and interpretability)

To support interpretability of the computational parameters used for clustering, we evaluated parameter recoverability via a single simulation-fitting pass: synthetic data were generated from the hierarchically fitted group posteriors and refitted with the same model, and agreement between generative and recovered parameters was quantified using ICC (Table 4). *ICC* values are reported both jointly across datasets and separately for each dataset. Most parameters showed good recovery. The inverse temperature parameter *β* was an exception, showing poor recoverability across datasets, suggesting weak identifiability for this quantity under the present task/model.

Parameter estimates were stable across random initializations, with low standard errors for cluster-level parameters across the joint fit (Supporting Information 1.3.1; mean standard error (*SE*^*m*^) across parameters: Cluster 1: *SE*_*m*_ = .024, Cluster 2: *SE*^*m*^ = .025) and for each dataset separately (Dataset I—Cluster 1: *SE*^*m*^ = .009, Cluster 2: *SE*^*m*^ = .014; Dataset II—Cluster 1: *SE*^*m*^ = .225, Cluster 2: *SE*^*m*^= .028). The notably higher S*E*^*m*^ for Dataset II Cluster 1 is consistent with the lower recoverability of that cluster noted above.

To validate the parameters further, we assessed correlations between fitted computational parameters and task-derived behavioural measures pooled across both datasets and clusters (Figure 3-3). Correlation patterns were consistent across datasets and clusters. Notable patterns include strong correspondence between computational and behavioural escape/avoid bias indices, and systematic associations between the Q-decay parameter and performance and strategy measures (accuracy and win–stay negatively; lose–switch positively).

##### Interpretative caveats (low *β* recoverability)

A key caveat concerns the inverse decision temperature parameter *β*, which could only be poorly recovered. Nevertheless, there are two reasons for continuing to interpret *β* (albeit cautiously). First, across both datasets, *β* exhibited highly consistent associations with model-agnostic behavioural features (Lose-Switch and Win-Stay), suggesting that it captures stable, behaviourally relevant variance in the fitted data even when absolute recovery is poor. In general, noisy parameter recovery would attenuate between-group differences (56), but in the current analyses, group differences remain detectable even after attenuation. Additionally, the low recoverability of *β* occurs in isolation, meaning all other model parameters are moderately to well recovered and showed very low covariance of *β* with other computational parameters (reported in Supporting Information Figure 1-1), which argues against *β* merely absorbing variance from learning-rate or bias parameters. Accordingly, we treat *β* as an interpretable index of choice stochasticity in this paradigm, whilst emphasising that inferences involving *β* should be considered provisional and evaluated alongside convergent task-derived signatures.

**Figure 4-3.**
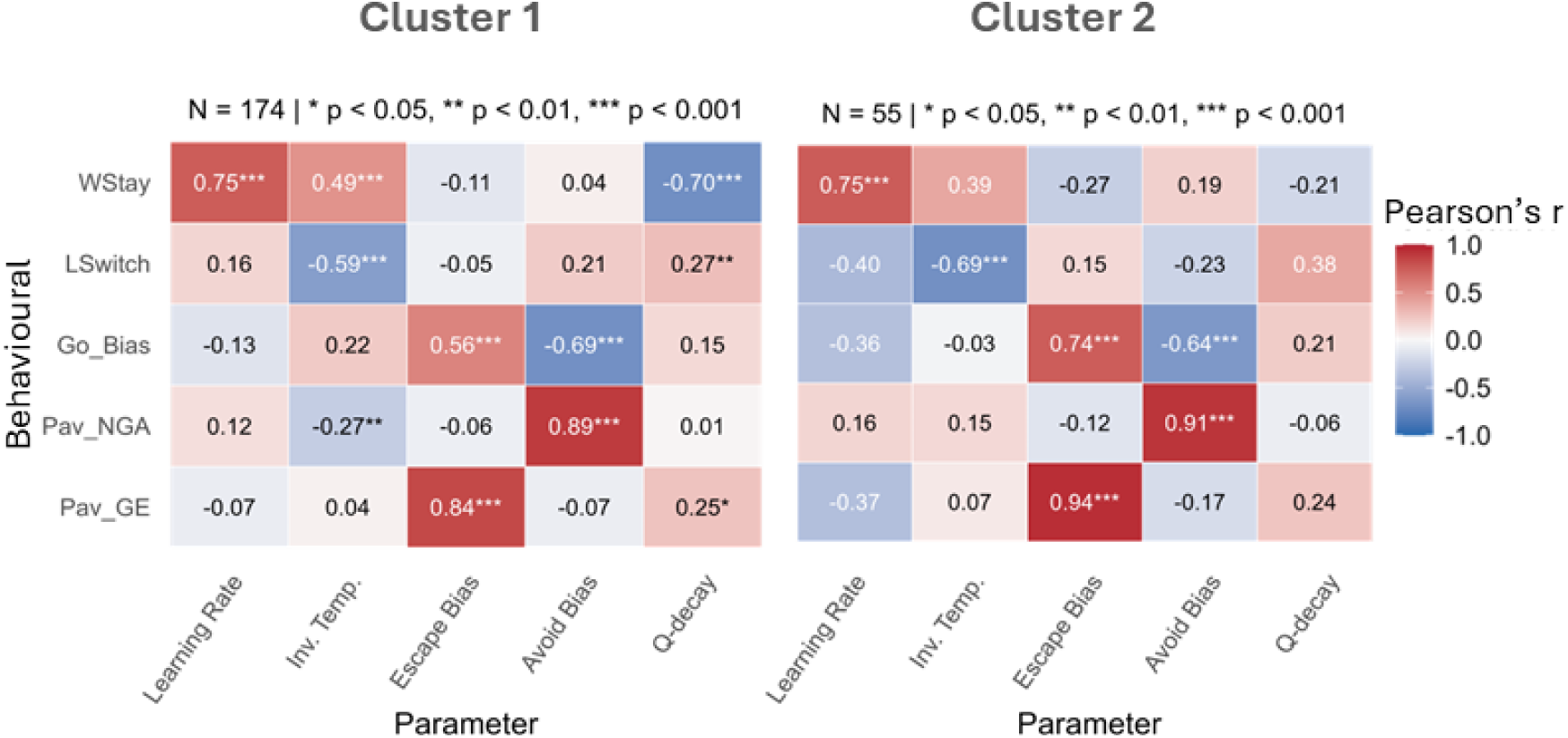
Parameter interpretability of computational parameters. Heatmap of Pearson correlations between fitted computational parameters and task-derived behavioural metrics, shown at the population level (combined fit). The pattern of associations establishes that parameters capture distinct and behaviourally interpretable aspects of task performance. GE = Go-to-Escape; NGA = NoGo-to-Avoid; LSwitch = Lose–Switch; WStay = Win–Stay, * indicates p<0.05, ** indicates p<.01, *** indicates p<.001 after Bonferroni correction.

### 4.3 Regime Characterisation: Computational and Behavioural Signatures

Across both datasets, the eHBI procedure identified heterogeneity in decision strategies. In both datasets, a two-cluster solution recurred across random initializations and yielded behaviourally distinct groups characterised primarily by differences in learning dynamics and choice policies. Below, we summarise each dataset’s clustering solution in terms of fitted parameter profiles, task-derived behavioural signatures, and model fit, and then evaluate whether stratification yields more specific clinical–computational couplings than pooled population analyses.

#### 4.3.1 Cluster Classification

##### Model-dependent subgrouping and behavioural signatures

In the joint fit, Cluster 1 comprised 174 participants and Cluster 2 comprised 55, with 5 participants unassigned due to insufficient label stability across initialisations (≥6/8 runs criterion). Cluster 1 consisted of 29 subjects from dataset I and 145 subjects from dataset II, while Cluster 2 consisted of 21 subjects from dataset I and 34 subjects from dataset II. We refer to Cluster 1 as the fast/adaptive regime and Cluster 2 as the slow/perseverative regime. These labels are motivated by the computational signatures described below: the fast/adaptive regime is characterised by rapid policy updating, high feedback reactivity, and relatively fast value decay, while the slow/perseverative regime is characterised by slow belief updating, high choice determinism, and persistent value retention that incurs a pronounced cost when previously learned contingencies are changed.

###### Computational parameter profiles

Across computational parameters, clusters differed significantly on four of five parameters (Bonferroni-corrected two-sided *t*-tests and Cohen’s d* based on the non-pooled standard deviation; Fig. 3-4). The slow/perseverative regime showed substantially higher choice determinism (β; *p_bonf_* <.001, *d\** = −4.14), consistent with committed, perseverative responding rather than exploratory choice. Its lower learning rates (*p_bonf_* <.001, *d\** = -1.37) indicate that outcomes exert less trial-by-trial influence on policy revision, while lower Q-decay (*p_bonf_* = 2.78e⁻², *d\** = 0.34) means that previously acquired values accumulate and are retained rather than being gradually reset. Together, these parameters instantiate a control strategy in which learned action preferences are slow to form but also slow to dissolve, a computational signature of perseveration at the level of value representation.

The fast/adaptive regime, by contrast, combined faster learning with higher value decay and greater choice stochasticity, a profile consistent with a volatile, feedback-driven control strategy in which policy representations are both rapidly updated and more rapidly abandoned.

**Figure 4-4.**
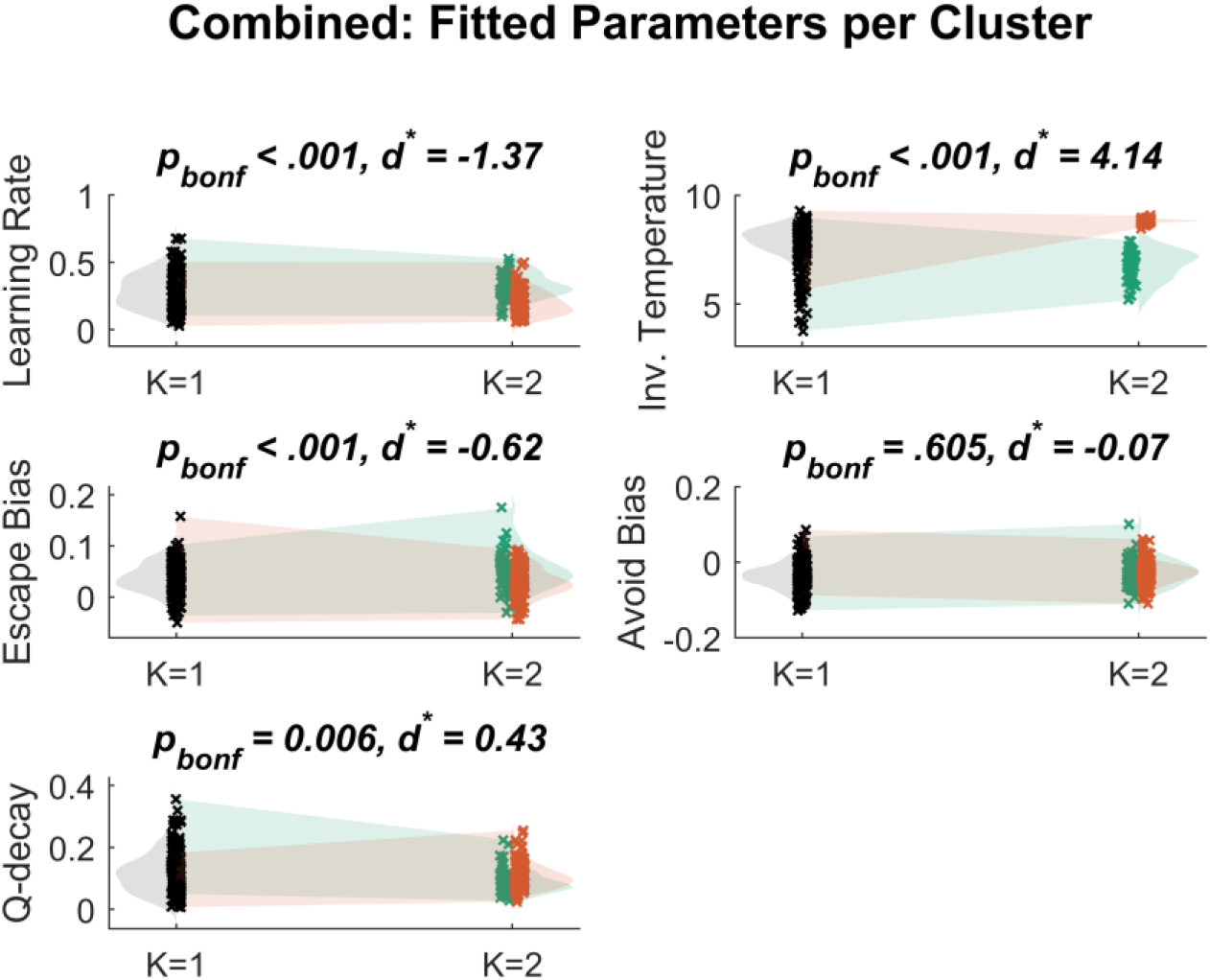
Fitted computational parameters across clusters. Plots show the posterior parameter distributions when data are fitted as a single population (K=1, grey) compared to the two-cluster solution (K=2, coloured by cluster, green- Cluster 1: fast/adaptive, orange – Cluster 2: slow/perseverative’). Parameters are shown for the joint fit across both datasets. Clusters are distinguished most clearly by Learning Rate, Inverse Temperature (β), and Escape Bias; Avoid Bias shows minimal separation. *p*_*bonf*_: Bonferroni corrected p-values, d*: Cohen’s d* based on the non-pooled standard deviation (57)

###### Behavioural signatures

Behaviourally, the clusters diverged on several task-derived measures (Fig. 3-5). The fast/adaptive regime showed substantially higher lose-switch behaviour (*pbonf* = 1.86e⁻¹⁵, *d* = 1.371), consistent with stronger sensitivity to recent aversive feedback. Pre-reversal accuracy was higher in the slow/perseverative regime (*pbonf* = 5.31e⁻¹¹, *d* = −1.12), while post-reversal accuracy was modestly higher in the fast/adaptive regime (*pbonf* = 2.95e⁻², *d* = 0.454), reflecting the cost of contingency change for the slow/perseverative cluster. The fast/adaptive regime also showed a higher overall Go rate (*pbonf* = 1.69e⁻², *d* = 0.481) and lower Pavlovian NoGo-to-Avoid index (*pbonf* = 1.45e⁻², *d* = −0.488).

**Figure 4-5.**
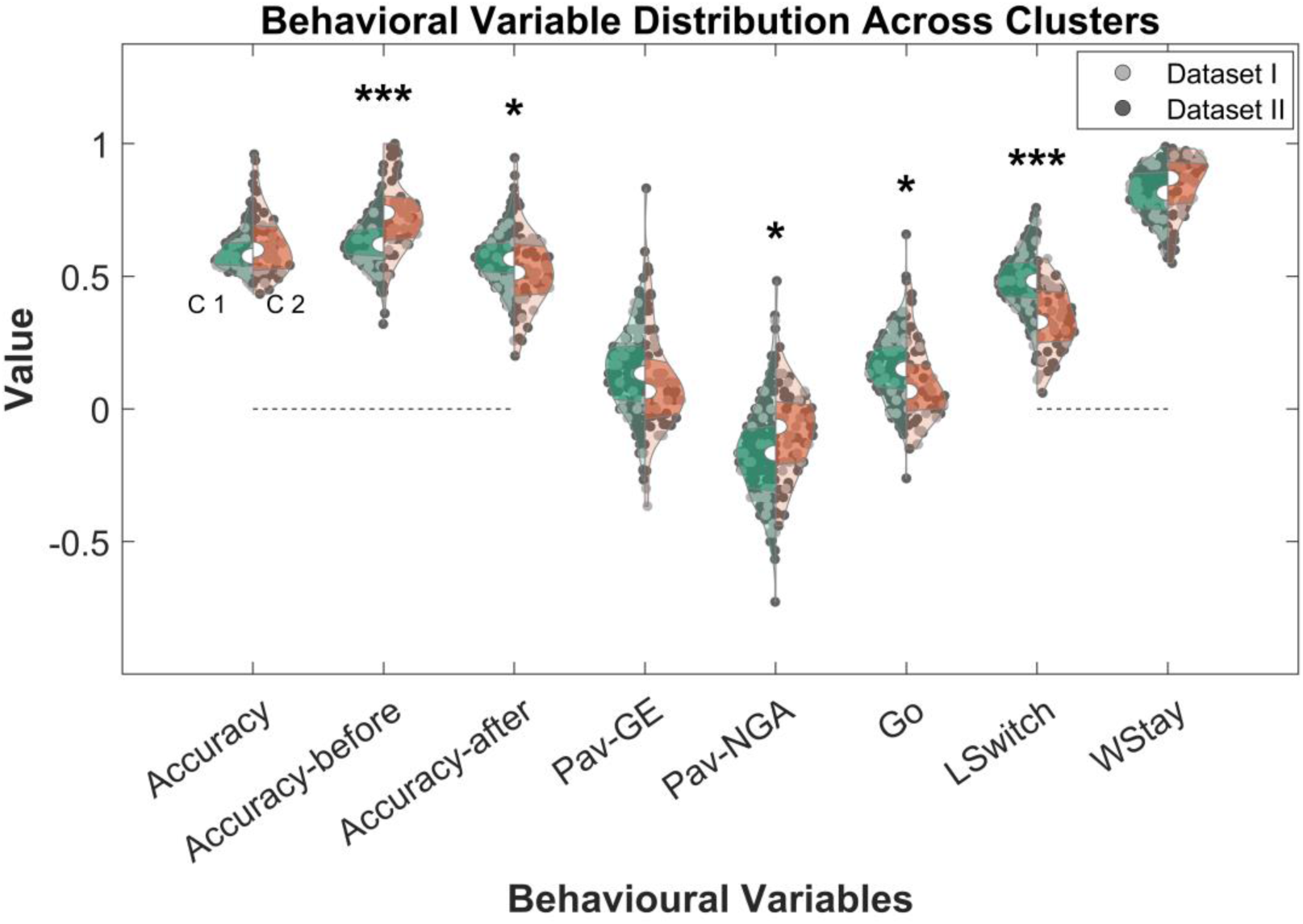
Task-derived behavioural differences across clusters. Violin plots show the distribution of behavioural measures for each cluster (Cluster 1: fast/adaptive, Cluster 2: slow/perseverative), with an overlaid split by dataset to confirm that cluster separation is not driven by dataset membership. Dotted lines at x=0 indicate measures that cannot be smaller than zero. Clusters differed significantly in lose-switch rate, pre- and post-reversal accuracy, overall Go rate, and Pavlovian NoGo-to-Avoid index; statistical comparisons after Bonferroni correction are reported in the main text. GE = Go-to-Escape; NGA = NoGo-to-Avoid; LSwitch = Lose–Switch; WStay = Win–Stay.

Trial-wise behavioural trajectories further characterised these regime differences (Fig. 3-6). In the fast/adaptive regime, accuracy during the pre-reversal phase was marked by greater short-term fluctuations following feedback, consistent with rapid policy updating. Following the contingency reversal, accuracy recovered relatively quickly in this regime. In the slow/perseverative regime, pre-reversal accuracy was higher overall, with a clearer early learning curve and smaller feedback-driven oscillations. Following reversal, however, this regime showed a pronounced accuracy drop with slower recovery, reflecting the difficulty of adapting when previously learned contingencies changed. Across both regimes, the RL-Pf model reproduced the overall trajectory patterns but showed systematic deviations in the 50 trials following reversal, where it over- or underestimated empirical performance. In-sample next-choice prediction accuracy was higher in the slow/perseverative regime than the fast/adaptive regime (Cluster 1: 0.67 ± 0.001; Cluster 2: 0.76 ± 0.002).

As expected, given that clinical variables were not included in the clustering procedure, regime assignments showed no significant differences in assessed clinical or demographic characteristics at the group mean level (Supporting Information, Figure 1-3). This confirms that the identified regimes reflect behavioural control strategies rather than clinical severity strata, and that any downstream clinical–computational associations are not trivially explained by cluster-level clinical confounds.

**Figure 4-6.**
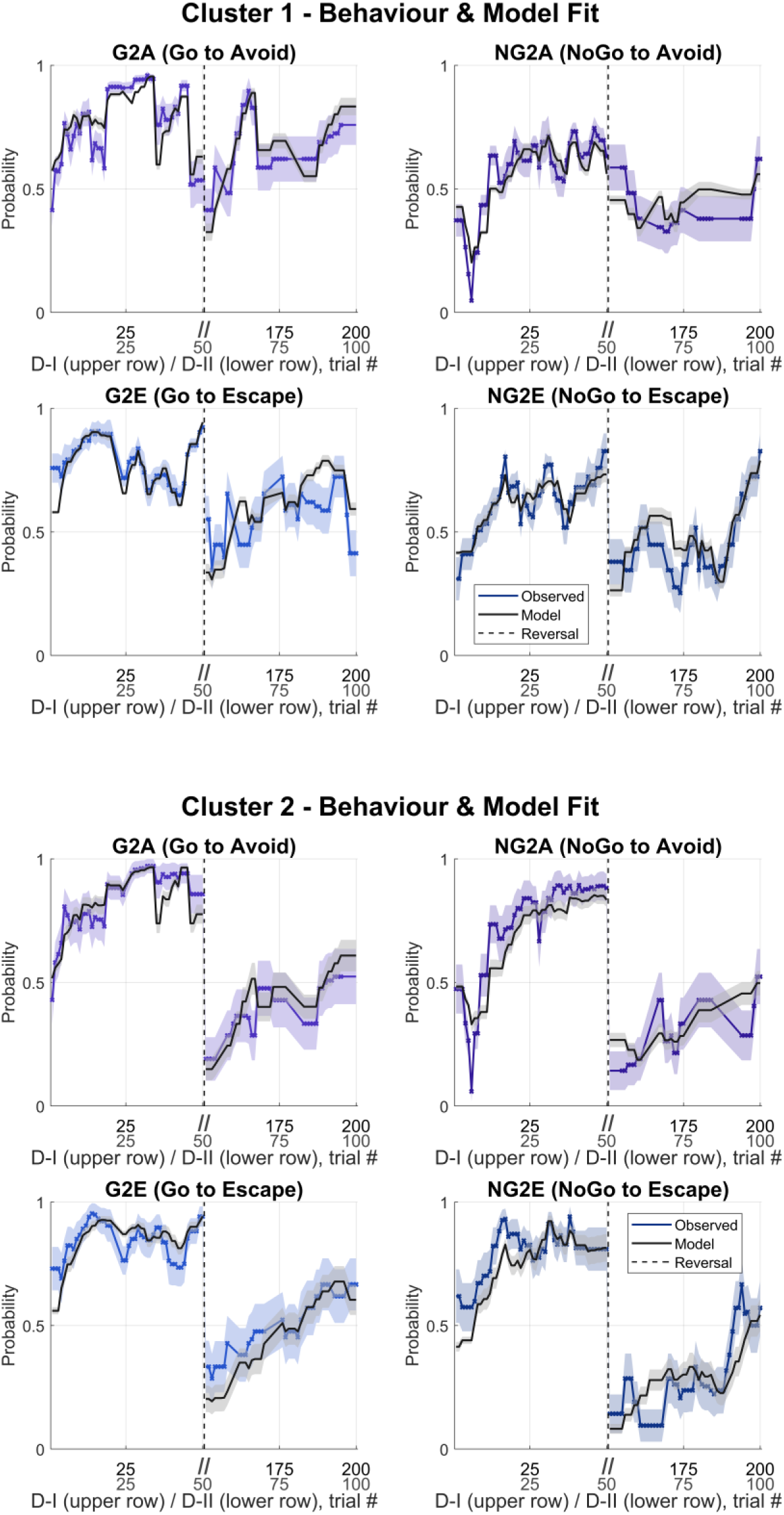
Behavioural trajectories and model fit split by cluster. Empirical trial-wise accuracy (coloured) and model-predicted accuracy (grey) are shown for the first 50 trials of the task and the first 50 trials immediately following the contingency reversal, separately for Cluster 1 (fast/adaptive, left panel) and Cluster 2 (slow/perseverative, right panel), pooled across datasets. Shaded regions indicate the 95% confidence interval. The two clusters diverge most clearly in the post-reversal period, with the slow/perseverative regime showing a more pronounced accuracy cost following the contingency change. D-I: Dataset I, D-II: Dataset II

#### 4.3.2 Comparing clinical to model-driven clustering

Having identified behavioural regimes through model-dependent clustering, we next examined whether these regimes correspond to patterns captured by conventional symptom-based stratification. To this end, participants were clustered using k-means based solely on clinical variables, and the resulting clinical groups were compared to the computational clusters identified above (see Methods for details).

##### Dataset I

Clinical clustering identified two groups of similar sizes (n = 26 and 24). Parameter comparisons again revealed minimal computational differences between clinically derived clusters. Significant differences were observed in inverse temperature (β) (*p_bonf_* = 5.06e⁻⁴, *d* = −0.456) and avoid bias (*p_bonf_* = 9.29e⁻⁴, *d* = 0.436), whereas learning rate, escape bias, and Q-decay did not differ significantly after correction.

The distribution of computational regime assignments was similar across the two clinical clusters: clinical cluster 1 comprised 42.3% fast/adaptive and 57.7% slow/perseverative participants, while clinical cluster 2 comprised 58.3% fast/adaptive and 41.7% slow/perseverative participants- This near-baseline distribution indicates that clinical clustering did not preferentially recover the computational regimes.

##### Dataset II

Clinical clustering identified two groups (*n* = 71 and 113). As in the other datasets, parameter differences between clusters were largely absent. The only strong difference was observed in inverse temperature, (*p_bonf_* < .001, *d* = −1.520), whereas learning rate, escape bias, avoid bias, and Q-decay did not differ significantly after correction.

Clinical cluster 1 comprised 80.0% fast/adaptive and 16.9% slow/perseverative participants; clinical cluster 2 comprised 77.9% fast/adaptive and 19.5% slow/perseverative participants. The full-dataset prevalence of fast/adaptive 78.9%. Clinical stratification did not reproduce the computational regimes.

Across both datasets, clustering based solely on clinical variables did not recover the computational regimes. Differences between clinically derived clusters were largely restricted to β – a parameter reflecting choice stochasticity rather than the learning and Pavlovian mechanisms central to the regime distinction. The distribution of computational regimes remained approximately constant across clinical clusters, confirming that the computational stratification cuts across conventional symptom-based groupings. Two individuals may appear clinically similar while relying on fundamentally different control strategies under aversive uncertainty, and this is precisely where computational subtyping adds value, not by replacing symptom characterisation but by providing a mechanistic layer that is sensitive to a different dimension of individual variation.

#### 4.3.3 Clinical–computational coupling

##### CCA overview

The joint CCA identifies canonical variates — linear combinations of variables — that maximally link the computational parameter space to the clinical variable space across both datasets simultaneously. Since the analysis is joint, the computational weight vector is shared; however, because the clinical instruments differ across datasets, the clinical weight vectors are estimated separately. Results are reported at two levels: across all participants (population level) and separately within each behavioural regime (cluster level). Loadings reflect the correlation of each variable with its canonical variate; only the pattern of relative loadings is interpretable, because the absolute magnitude of a canonical variate is arbitrary.

As noted, the two datasets differ substantially in clinical instrumentation — ACE, BDI-II, BAI-T, STHS, BIS-11, BPAQ, and GAD-7 in Dataset I versus HiTOP-SR, INQ, and GCSQ in Dataset II — as well as in recruitment context. Apparent divergences in clinical loading patterns across datasets may therefore reflect differences in instrument coverage rather than genuine heterogeneity in the underlying computational–clinical relationships. The shared computational parameter space, identical across datasets by design, provides the common axis along which cross-dataset consistency can be assessed; convergence is examined in Section 3.3.2.

##### Dataset I (reversal, 300 trials)

###### Population-level clinical–computational relationships

The population-level axis links an escape-biased, stochastic Pavlovian control profile to a broad pattern of concurrent affective burden and active suicidality, consistent with a general psychopathology dimension rather than a suicidality-specific one. In the statistically significant first canonical covariate (CV; *r* = .319, *p* = .045, Fig. 3-7), the computational axis was dominated by Escape Bias with a secondary contribution from Inverse Temperature and Avoid Bias, reflecting a Pavlovian action-bias dimension in which higher escape bias and greater choice stochasticity co-occurred with lower avoidance bias. Clinically, higher escape-bias co-occurred most strongly with depression, anxiety (BAI-T, GAD-7), hopelessness, and recent suicidal ideation severity, while suicidal behaviour, aggression and impulsivity had more moderate weights. Notably, recent suicidal ideation severity weighted substantially more strongly than lifetime ideation history, suggesting this axis captures acute psychopathology rather than historical burden.

**Figure 4-7.**
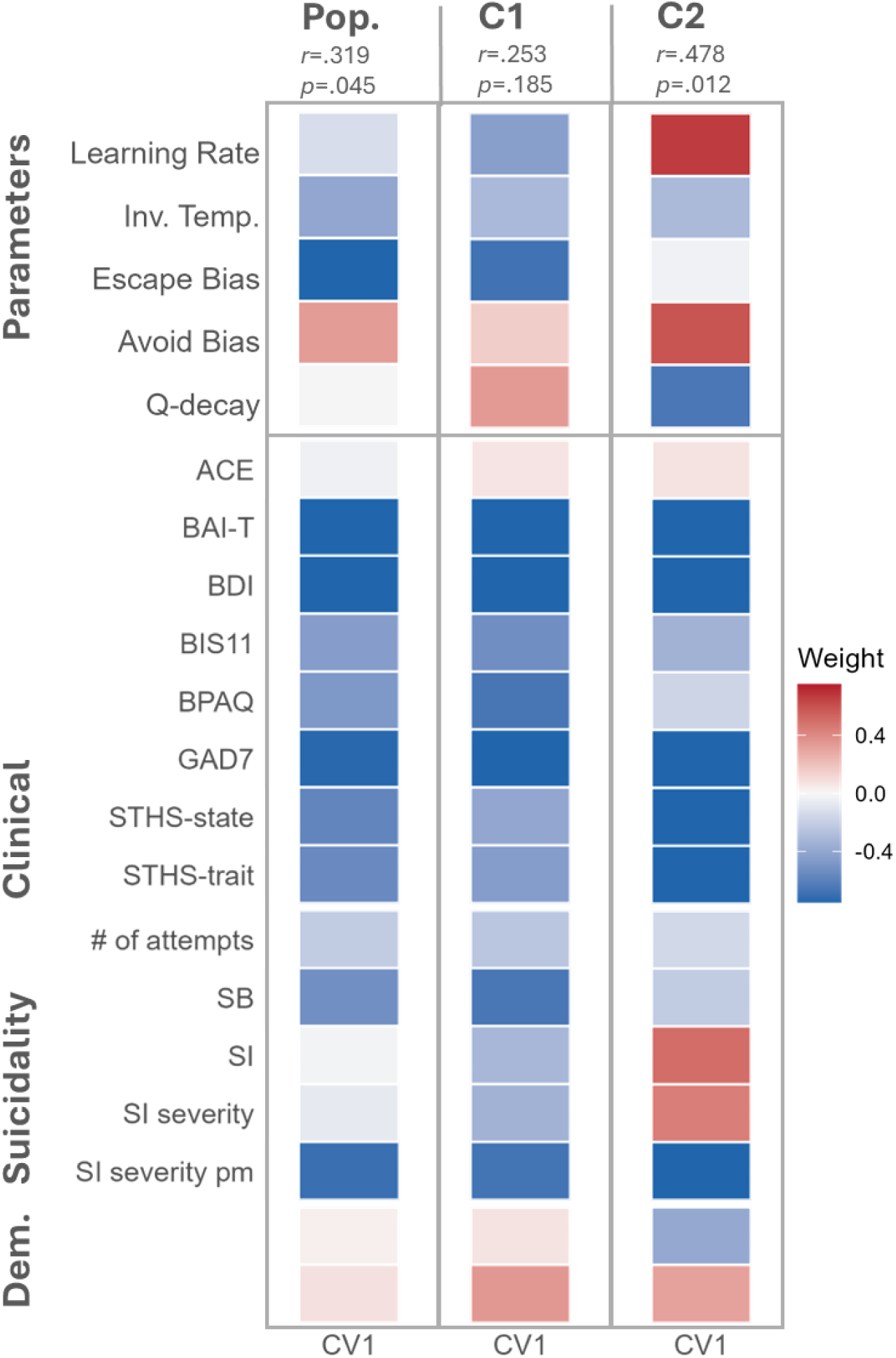
Canonical correlation analysis results for Dataset I, showing the first canonical variate for each CCA. Heatmaps of weights, showing the weight of each canonical variate onto computational parameters (top) and clinical/suicidal variables (bottom). More intense colours correspond to higher absolute weights. At the population level, the dominant axis links an escape-biased Pavlovian control profile to broad affective burden. In Cluster 1, the axis reflects the population pattern but does not retain statistical significance. In Cluster 2 (slow/perseverative), the axis shifts to learning patterns, with a stronger and more specific negative association to internalizing burden. MDD: major depressive disorder, C-SSRS: Columbia Suicide Severity Rating Scale, BDI-II: Becks Depression Inventory, BIS11- Barrat Impulsiveness Scale, GAD7: Generalized Anxiety Disorder Scale-7, ACE: Adverse Childhood Experience, STHS: State-Trait Hopelessness Scale, BPAQ: Buss Perry Aggression Questionnaire, BAI-T: Beck’s Anxiety Inventory-Trait, SB: Suicidal Behaviour, SI: Suicidal Ideation, Dem: Demographic

###### Cluster-level clinical–computational relationships

Repeating the joint CCA within each cluster yielded more differentiated coupling patterns than at the population level, consistent with mixture cancellation masking regime-specific associations in the pooled sample. Given modest cluster sample sizes, these results should be treated as exploratory.

*Fast/adaptive regime* (Cluster 1): The single extracted canonical variate did not reach statistical significance (*r* = .253, *p* = .185). Its structure mirrored the population-level axis, namely an escape-biased Pavlovian control profile co-occurring with broad concurrent affective burden, suggesting that within this regime the population-level signal is preserved but not sharpened by stratification.

*Slow/perseverative regime* (Cluster 2): Stratification produced a more pronounced clinical-computational structure, where the accuracy-linked computational axis was associated with lower current affective burden alongside an increased history of elevated lifetime ideation. The coupling observed was the strongest in cluster 2 (*r* = .478, *p* = .012). The computational axis shifted away from Pavlovian escape bias entirely – e.g., Escape Bias dropped to near-zero - and was instead defined by Learning Rate, Q-decay, and Avoid Bias, a profile in which faster learning, lower value decay, and stronger avoidance bias co-occurred with higher task accuracy (*r* = 0.72, *p* = 0.0003 with canonical variate). The pattern of relationships among clinical variables was preserved relative to the population level but emphasized more specific anchors, such as anxiety (BAI-T, GAD-7), hopelessness, depression, and recent suicidal ideation severity all weighted strongly, while aggression and impulsivity (i.e., externalizing problems) were markedly attenuated. The dissociation of lifetime ideation severity loading in the opposite direction as recent ideation severity and current affective burden did not replicate in bootstrapping analyses, as recent ideation did not emerge as a significant contributor to the CV. Weight on demographic variables is increased compared to the population level, but clinical correlates remain stronger contributors to the CV axis (age: -.39, sex:.31).

##### Dataset II (120 trials, reversal)

###### Population-level clinical–computational relationships

At the population level, we observed a correlation of Pavlovian action-bias structure with a strong association of INQ and relatively higher psychopathological burden (low overall weight, Fig. 3-8). The only significant canonical variate (CV1: *r* = .214, *p* = 8.92e⁻⁶) produced a computational axis dominated by Escape Bias and Inverse Temperature, with Avoid Bias contributing positively. Higher escape bias and greater choice stochasticity negatively co-occurred most strongly with unmet interpersonal needs as measured by the INQ, capturing perceived burdensomeness and thwarted belongingness, alongside NSSI (a sub-item of HiTOP), antisocial/disinhibitory behaviour, and internalizing distress features including depressed mood and anhedonia. In contrast to Dataset I’s population axis, which was anchored by depression and anxiety, the primary clinical anchor here is interpersonal suicidality embedded within a mixed internalizing–externalizing profile, with perceived capability for suicide dissociating from affective burden.

Despite these population-level associations, clusters did not differ significantly in clinical measures (HiTOP internalizing/externalizing spectra, INQ, GCSQ) or in demographic characteristics at the cluster-mean level. Note that we combined the INQ dimensions of thwarted belongingness and perceived burdensomeness to increase readability, as weights across CCAs were almost identical in all cases.

**Figure 4-8.**
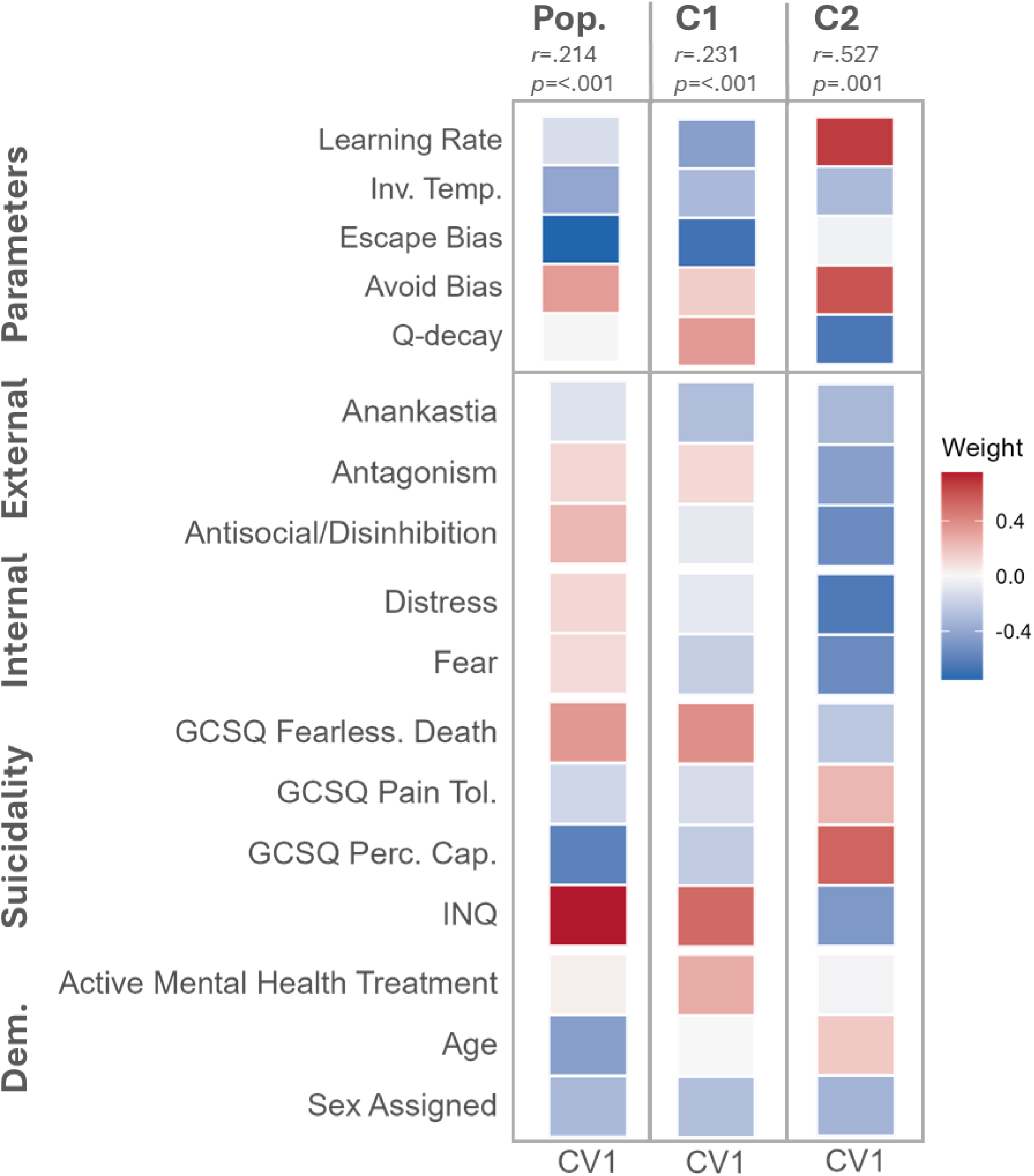
Canonical correlation analysis results for Dataset II. Heatmaps of weights, showing the weight of each canonical variate onto computational parameter (top) and HiTOP/suicidal variable (bottom) at population and cluster level. More intense colours correspond to higher absolute weights. At the population level, CV1 links an escape-biased computational profile to interpersonal suicidality (INQ) and mixed internalizing–externalizing burden; CV2 links a learning-efficiency axis to cognitive-executive dysregulation. Cluster 2 (slow/perseverative) yields the largest effect size across all analyses, with the avoid-bias/learning-pattern axis associating broadly with lower psychopathology and a dissociation between distress measures and GCSQ Perceived Capability., INQ: Interpersonal Needs Questionnaire, GCSQ: German Capability for Suicide, Dem: Demographic

###### Cluster-level clinical–computational relationships

Within-cluster analyses in Dataset II showed the same pattern of amplified and reorganised coupling, with effect sizes exceeding those at the population level in both regimes.

*Fast/adaptive regime*: The canonical variate for Cluster 1 separates an anxious/compulsive profile from an antisocial/disinhibited/NSSI profile along a computational axis of Pavlovian escape and adaptive learning. The significant CV (*r* = .231, *p* = 7.10 × 10⁻⁷), reproduced the escape-biased, adaptive profile seen at the population level – Escape Bias, Learning Rate, and Q-decay – but stratification revealed an internal clinical dissociation that was obscured in the pooled sample. Note that while the learning rate weight was stable, the weight on Escape Bias in CV1 did not hold up under bootstrapping analysis. At one pole, higher escape bias and faster reactive learning co-occurred with anxious features (checking, hoarding, anxious worry) and consistent anankastia items (perfectionism, risk aversion, etc.) alongside cognitive disorganization. This combination may reflect the interference of anxious preoccupation with executive function rather than a clean compulsive profile. At the opposite pole, lower escape bias and slower, more stochastic learning co-occurred with more antisocial behaviour, callousness, NSSI, and unmet interpersonal needs. This within-cluster dissociation was not visible at the population level and represents a clinically meaningful stratification not recoverable from pooled analysis. The behavioural association should be reproduced in larger samples, as bootstrapping revealed variability in the implicated parameters.

*Slow/perseverative regime*: In this regime, a higher avoidance bias and more efficient learning profile co-occurred with broad reduction in psychopathology and increased perceived capability, while the opposite pole with decreased learning and value retention efficiency was associated with increased affective burden and unmet interpersonal needs. The significant CV (*r* = .527, *p* = .0014), yielded the largest effect size across all analyses in either dataset. The computational axis was dominated by Avoid Bias, Learning Rate, and Q-decay, consistent with the learning-dynamics structure observed in Dataset I’s Cluster 2, and was weakly positively correlated with task accuracy before reversal (effect did not survive Bonferroni correction, see SI 1.4.4.3). On the clinical side, we observed a reduction in psychopathology across four clinical domains: externalizing antagonism (dishonesty/deceitfulness, manipulativeness, entitlement, social aggression); externalizing antisocial (disorganization, nonplanfulness, nonpersistence); internalizing distress (anxious worry, shame/guilt, irritability, depressed mood); and internalizing fear (hoarding, checking). Unmet interpersonal needs (INQ) loaded in the same direction. The single prominent exception was GCSQ Perceived Capability, which ran in the opposite direction to virtually all other clinical variables including INQ. Within this regime, perceived capability for suicide thus dissociates from the broader psychopathology space along the learning-dynamics axis — aligning with the lower-avoidance, less efficient computational profile independently of general distress or ideation.

The contribution of demographic variables is small across all CCA analyses.

## 5 Discussion

Across two STB-enriched adult samples, joint computational modelling of aversive Go/NoGo behaviour identified two recurrent behavioural regimes and revealed that stratification by regime exposes clinical–computational coupling structures that are substantially attenuated in pooled analyses. Three findings address the pre-specified questions directly.

First, the joint modelling identified two stable and behaviourally distinct regimes: (i) a fast/adaptive regime characterised by elevated lose-switch behaviour, stronger Go tendencies, faster policy updating, and greater sensitivity to recent feedback, and (ii) a slow/perseverative regime characterised by slower updating, more deterministic choice, and a pronounced performance cost when previously learned contingencies changed, indicating stronger reliance on retained value representations and reduced flexibility under environmental change. The recovery of cluster assignments across datasets differing substantially in task length, reversal timing, and sample composition, indexed by above-chance agreement in joint modelling, suggests that these regimes reflect a shared computational structure rather than dataset-specific artefact (RQ1).

Second, these regimes were largely orthogonal to symptom-based stratification: clinical clustering did not reproduce the computational subtypes, and cluster means on clinical variables were similar across regimes. This dissociation is consistent with the expectation that cross-sectional symptom scores are insensitive to the trial-level behavioural dynamics the task directly measures, focusing on how rapidly individuals update policies following feedback, how strongly recent outcomes drive the next choice, and how quickly previously learned values decay — trial-level properties with no direct counterpart in a single-timepoint questionnaire (RQ2). Consistent with prior mechanistic suggestions about suicidal subtypes(22), the regimes diverged most clearly in learning dynamics, feedback reactivity, and action bias structure rather than in overall task performance or clinical severity.

Third, and most centrally, stratification by regime produced markedly differentiated clinical–computational coupling (RQ3). We started at the population level, where canonical associations between computational parameters and clinical variables were statistically significant but modest across both datasets. Here, the dominant axis was anchored by Pavlovian escape bias and linked to broad affective burden and current suicidality. This is a general psychopathology dimension rather than a suicidality-specific one: it reflects the well-established association between aversive reactivity and clinical distress that would be expected in any sample enriched for STBs, and its modest effect size is consistent with the expectation that opposing regime-specific couplings attenuate each other when participants are pooled.

However, individuals may appear clinically similar while relying on fundamentally different control strategies under aversive uncertainty — and this is precisely where computational subtyping may add value, not by replacing symptom characterisation but by providing a mechanistic layer that is sensitive to a different dimension of individual variation. Thus, within the slow/perseverative regime, effect sizes were substantially larger, the computational axis shifted from a Pavlovian bias to learning dynamics and avoidance, and the clinical structure reorganised around internalizing burden rather than broad psychopathology. If the slow/perseverative regime’s value-retention profile is mechanistically upstream of the hopelessness and sustained ideation that define its internalizing clinical signature, then the computational parameters are not merely correlates of that signature but candidate generative causes. The present data are cross-sectional and cannot establish this directionality, but they motivate a programme in which computational task design is guided by the clinical heterogeneity one seeks to explain.

In reinforcement learning terms, the within-regime gradient in the slow/perseverative cluster tells a specific mechanistic story. At the higher-burden and unmet interpersonal needs pole (higher Q-decay and lower learning rate) outcomes fail to drive targeted value revision, and the value landscape erodes passively with each passing trial regardless of what occurs. These are two distinct failures: low learning rate reflects reduced sensitivity to specific outcome contingencies; high decay reflects indiscriminate erosion of all value representations independent of feedback. Together they describe a system that neither learns efficiently from experience nor maintains stable representations of what it has learned; disengagement from environmental contingency rather than rigid value retention. The lower-burden pole with increased suicidal capability scores, and computationally higher learning rate and lower decay, learns more effectively and maintains more stable representations, consistent with its higher pre-reversal accuracy.

This pattern resonates with converging evidence linking reduced aversive learning rates to internalizing burden in depression (58), reduced ability to modulate behaviour as a function of reinforcement history to motivational withdrawal and anhedonic symptoms (59), and perceived uncontrollability of aversive outcomes to hopelessness through suppression of effective aversive learning (22). Within the slow/perseverative regime, those for whom aversive contingencies are least informative and least stably represented carry the greatest internalizing burden — a profile in which hopelessness and sustained ideation may reflect the progressive decoupling of action from meaningful environmental feedback.

Within the fast/adaptive regime, Dataset II resolved an internal dissociation between anxious–compulsive and antisocial–disinhibitory profiles along the same computational axis, a distinction not visible at the population level. Further, the coupling pattern tells a different story from the slow/perseverative regime. Associations were weaker overall, and the computational axis retained the same population-level structure, suggesting that within this regime the mapping between computational parameters and clinical burden is less differentiated than at the population level.

The one exception to this lack of differentiation, visible only in Dataset II with its different clinical instrumentation, is the internal dissociation between anxious–compulsive and antisocial–disinhibitory profiles along the same computational axis. This dissociation suggests that within the fast/adaptive regime, suicidality-relevant clinical variance may be distributed across internalizing and externalizing pathways that happen to sit at opposite poles of the same computational dimension, a structure that may not be detectable in pooled analyses. This pattern parallels the transdiagnostic compulsivity/impulsivity distinction identified in large online samples, where compulsivity and disinhibition emerge as separable computational phenotypes that are obscured within conventional diagnostic groupings (65,66). The behavioural profile of the fast/adaptive regime – faster policy updating, elevated lose-switch behaviour, higher value decay, and stronger action tendencies – is structurally consistent with prior descriptions of stress-reactive or temporally variable forms of suicidal risk, though the clinical data from the present study do not directly support this mapping. The present datasets lack longitudinal ideation monitoring and stress-exposure measures, which are the instruments that would be needed to test this correspondence directly. Studies of ideation dynamics are relevant as a behavioural parallel: (67) and (15) identified phenotypes defined by frequency, intensity, and variability, with high-variability groups showing elevated odds of subsequent attempt. The fast/adaptive regime shares a structural resemblance with these temporally unstable profiles — both implicate rapid fluctuation in response to environmental input rather than stable, persistent states. Clinical subgrouping studies describing externalizing or impulsive subgroups (68,69) and stress-reactive phenotypes (70,71), alongside biological evidence linking HPA-axis reactivity to impulsive-aggressive attempter subgroups (16,17), converge conceptually on this profile.

Finally, fearlessness-of-death co-varied with antagonistic and disinhibitory traits rather than with internalizing burden along the same computational axis. This directional pattern aligned with an action-oriented, externalizing profile rather than a distress-driven one, but it should be treated as hypothesis-generating given the exploratory nature of the cluster-level CCA and the modest sample sizes involved.

Taken together, the two regimes reveal qualitatively different kinds of clinical–computational structure, both invisible at the population level: the slow/perseverative signal is masked by mixture cancellation, while the fast/adaptive dissociation requires instrument sensitivity (e.g., HiTOP) that pooled analyses lack.

### Limitations and future directions

Several limitations constrain inference. Sample sizes, particularly for cluster-specific analyses, were modest, which limits precision and increases uncertainty in estimated coupling patterns. Cluster-specific CCA should accordingly be regarded as exploratory, and effect sizes may be inflated in smaller subgroups.

The inverse temperature parameter was harder to recover than other parameters, which may reflect a limitation of the current model architecture rather than a property of the data alone. Lose-switch behaviour — the most prominent behavioural discriminator between regimes in the model-agnostic analyses — is not explicitly parameterised in RL-Pf but must instead be absorbed by the interplay of learning rate, Q-decay, and inverse temperature. This indirect representation may contribute to the identifiability problems observed for β and suggests that future model development should explore explicit parameterisation of feedback-driven switching as a distinct mechanism.

The joint modelling approach enforces a shared computational structure across datasets – a deliberate design choice that supports cross-dataset comparison but also assumes that the tasks in both datasets measure the same cognitive processes; deviations from this assumption would bias rather than simply add noise to the estimated regime assignments. While the comparison of task-derived behaviours and computational parameters across datasets supports the design choice, subtle task differences including the length and gamification may impact the cognitive strategy individuals employ beyond the assessed dimensions.

The clinical instrumentation differs substantially between datasets, meaning that apparent divergences in clinical loading patterns cannot be straightforwardly attributed to genuine heterogeneity in the clinical–computational relationship rather than to instrument coverage. This is particularly relevant for the tentative parallel observed in Cluster 2 between lifetime suicidal ideation (Dataset I) and perceived capability for self-harm (Dataset II), where the available data do not permit a definitive interpretation.

More broadly, the study is cross-sectional and task specific. We did not measure attempt planning, lethality, or longitudinal ideation dynamics, and therefore cannot determine whether the identified regimes predict clinically important future outcomes. Future work should test these regimes prospectively in larger samples, examine demographic variables as moderators rather than covariates, and relate behavioural regimes to ecological momentary assessment of ideation dynamics and stress exposure over time. Multi-task designs would help determine whether the regimes generalise beyond this aversive Go/NoGo paradigm or reflect a more specific form of control under contingency reversal.

### Conclusion

Model-dependent joint stratification of aversive Go/NoGo behaviour identified two recurrent behavioural regimes in suicidality-relevant samples: a fast/adaptive regime marked by rapid updating, action bias, and strong feedback sensitivity, and a slow/perseverative regime marked by slower adaptation and greater contingency-change cost. These regimes were reproducible across datasets and largely orthogonal to symptom-derived stratification. Most importantly, stratification by regime exposed clinical–computational coupling structures that were substantially attenuated in pooled analyses: the same clinical landscape decomposed differently once behavioural control regime was considered, with the slow/perseverative regime yielding the strongest and most interpretable associations. The results support a view of suicidality heterogeneity in which clinically similar individuals may differ in the control strategies they recruit under aversive uncertainty, and in which this mechanistic variation is not captured by standard symptom measurement. Rather than treating heterogeneity as noise around a single suicidal phenotype, this approach treats it as structure that can be measured behaviourally and interpreted mechanistically. The next step is to test whether these regimes are reproducible in larger samples, prospectively informative, and clinically useful complements to symptom-based characterisation.

## Data Availability

All data produced in the present study are available upon reasonable request to the authors.

