## Supplemental Information for "Two modes of aversive control in suicidality: joint computational modelling exposes regime-specific clinical signatures invisible to symptom-based stratification"

### 1 Supporting Information

#### 1.1 Data

##### 1.1.1 Exclusion Criteria

Data exclusion criteria were based on accuracy and sequences of consistent Go or NoGo choices

Participants were excluded from subsequent analysis if they failed any of the following exclusion criteria.

1. Participants reached at least 55% average accuracy at least before or after the reversal.
2. Participants had an accuracy of less than 10% either before or after the reversal, indicating reversed association of action to cue
3. If a participant's longest sequence of Go or NoGo responses was outside of the population mean ± 3 standard deviations. For the evaluation, each participant's longest uninterrupted sequence of Go actions and their participants longest uninterrupted sequence of NoGo actions were identified.

The accuracy threshold was implemented as a data-quality criterion rather than as a subject-level inferential test. For Dataset I, we derived an a priori cut-off of 55% based on the exact binomial distribution under chance performance (n = 300 trials, pnull = 0.5), using a one-sided right-tail criterion with $\alpha = 0.05$.

Under this null model, 55% represents the lowest accuracy reliably exceeding chance and therefore provides a principled safeguard against near-random responding.

For consistency across datasets, we applied the same nominal 55% threshold to Dataset II. Due to the reduced task length, this corresponds to a one-sided binomial tail probability of approximately $\alpha = 0.16$ at 55% accuracy. The threshold was applied pre- and post-reversal to distinguish global disengagement from phase- or condition-specific variability in performance. This ensured that participants demonstrating above-chance performance in at least one task segment were retained.

##### 1.1.2 Task Design

*Table 1 Task Design details for aversive Go/NoGo tasks deployed in datasets I and II.*

| | Task – Dataset I | Task – Dataset I |
|---|---|---|
| Cue duration | 3s | 1-3s |
| Response Window | 3s | 3s |
| Feedback duration | 2s | 1-3s |
| Inter-trial duration | 0s (feedback served as inter-trial break) | 0s (feedback served as inter-trial break) |

##### 1.1.3 Task-derived measures

*Table 2 Model-agnostic behavioural measures across datasets. GE: Go-to-Escape Bias, NGA: NoGo-to-Avoid Bias*

| Measure | Dataset I | Dataset II |
|---|---|---|
| Overall Accuracy | 0.6828± 0.1062 | 0.5775± 0.0587 |
| Pre/post reversal accuracy | 0.7576± 0.1461<br><br>0.6063± 0.1340 | 0.6277± 0.0850<br><br>0.5417± 0.0967 |
| Go-Bias | 0.1584± 0.1788 | 0.1315± 0.1133 |
| Pav-GE | 0.2063± 0.2046 | 0.1072± 0.1477 |
| Pav-NGA | -0.1071± 0.2209 | -0.1558± 0.1626 |
| Lose-Switch | 0.4265± 0.1371 | 0.4483± 0.1242 |
| Win-Stay | 0.8135± 0.1056 | 0.8230± 0.0905 |

#### 1.2 Model selection

Model selection followed the empirical hierarchical Bayesian inference (eHBI) procedure (Laessing et al., 2025.; Piray et al., 2019), applied separately to each dataset. Candidate models explored mechanisms across 5 dimensions:

- Learning Mechanism represented by Reinforcement Learning (R), Kalman filter (K), Active Inference (AI)
- Pavlovian Mechanism in the form of context bias (b), evidence bias (a), policy (ω), policy with variable contribution (v)
- Global Go-bias (G)
- reward sensitivity by condition (ρ) influencing learning
- Mnemonic mechanism, including forgetting (f), working memory (m)

First, we systematically narrowed the model space to retain only those with significant evidence of capturing relevant mechanisms in the population (Fig 1-1). To this end, we evaluated individual model fits across each dataset, evaluating their fits based on model responsibilities, computed based on the individual fit quality, as captured by the likelihood,

$$r_k = \frac{\sum_{n=1}^{N} p(x_n | h_{n,k})}{\sum_{k=1}^{K} \sum_{n=1}^{N} p(x_n | h_{n,k})}$$

, where $h_{n,k}$ are the fitted parameters for the kth model and nth subject.

Across both datasets, the two Reinforcement Learning models with Pavlovian biases and mnemonic mechanisms performed best. In Dataset II, we additionally observed a 12% of individuals best fitted by the Active Inference model.

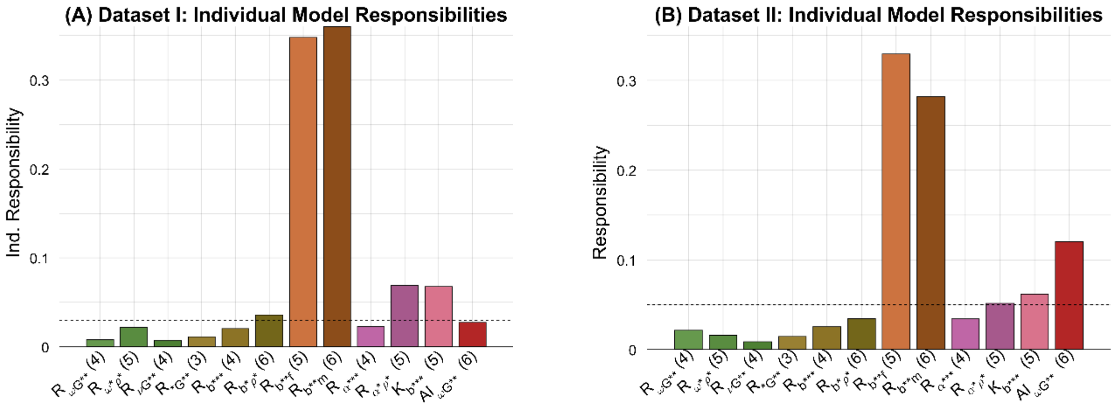

*Figure 1-1 Relative individual model responsibilities. Relative Model responsibilities associated with the full model space individually fitted to each individual in the full population for dataset I (A) and II (B), respectively. A total of 12 different models was fitted to each individual, using non-informative priors. The relative responsibilities were averaged across individuals to obtain a mean relative responsibility of each model for the studied populations. Dashed lines indicate the cut-off for each population to consider a model in the reduced model subspace. Thresholds were chosen empirically and adjusted to create a model space where the least prominent model would have a residual exceedance probability below .01. This approach was chosen to ensure a minimally biased fitting procedure, in which all models with the potential to explain a marked amount of behavioural variance were included. Note: Model notation reflects the different mechanisms as per the introduction. For ease of reference, we later refer to the winning model R_{b**f} as RL-Pf.*

Those models that scored above the responsibility threshold in each dataset were selected and compared with the empirical Hierarchical Bayesian Inference (eHBI) procedure. They were evaluated using model evidence and protected exceedance probability, which quantify integrated data fit and the posterior probability that a given model outperforms all alternatives, respectively (Fig 1-2).

The winning model (Reinforcement Learning with Pavlovian context bias and forgetting; RL-Pf) was identified consistently across both datasets with exceedance probabilities of $p_{e,I} = .98$, $p_{e,II} = 1$. The winning model was used for all subsequent analyses to discover clusters within the populations.

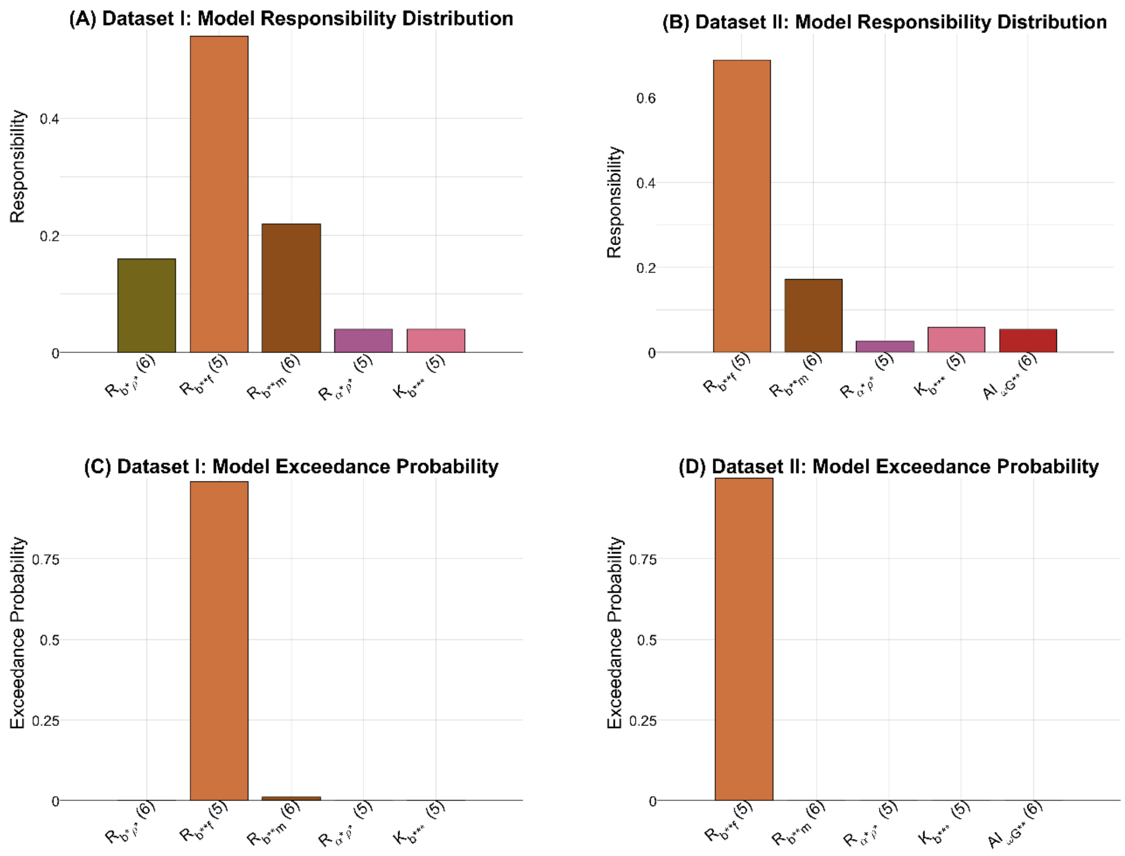

*Figure 1-2 eHBI model comparison results for dataset I and II. (A) and (B) show the responsibility distribution across the respective model subspaces. (C) and (D) show the exceedance probabilities for each model for dataset I and II, respectively. Results are consistent across both datasets, indicating the RL model with Pavlovian context bias parameters and a forgetting parameter clearly outperforms the competing models. Note: Model notation reflects the different mechanisms as per the introduction. For ease of reference, we later refer to the winning model R_{b**f} as RL-Pf.*

#### 1.3 Cluster selection

##### 1.3.1 Subgrouping comparison numbers

*Table 3. Model fit for configuration 2 for each dataset, including evidence lower bound, iBIC and predictive probability per choice (given individual group assignments). iBIC: integrated Bayesian information criterion, iAIC: integrated Akaike information criterion, EL: evidence lower bound, SE: standard error across runs*

| **Dataset I** | Config 1 | | | | Config 2 | | | | Config 3 | | | |
|---|---|---|---|---|---|---|---|---|---|---|---|---|
| | EL | iBIC | iAIC | P(choice) | EL | iBIC | iAIC | P(choice) | EL | iBIC | iAIC | P(choice) |
| Run 1 | -7070.50 | 14252.13 | 14168.45 | 0.70 | -7044.94 | 14304.29 | 14142.82 | 0.70 | -7037.21 | 14397.82 | 14127.53 | 0.70 |
| Run 2 | -7070.54 | 14260.95 | 14178.04 | 0.70 | -7044.19 | 14302.79 | 14131.22 | 0.70 | -7037.16 | 14396.48 | 14142.28 | 0.70 |
| Run 3 | -7070.40 | 14260.85 | 14165.28 | 0.70 | -7045.75 | 14302.49 | 14139.61 | 0.70 | -7038.04 | 14404.73 | 14133.74 | 0.70 |
| Run 4 | -7070.47 | 14250.47 | 14187.54 | 0.70 | -7043.55 | 14292.22 | 14136.29 | 0.70 | -7038.30 | 14394.00 | 14123.55 | 0.70 |
| Run 5 | -7070.40 | 14259.98 | 14172.45 | 0.70 | -7044.37 | 14309.75 | 14128.90 | 0.70 | -7038.63 | 14394.64 | 14133.73 | 0.70 |
| Run 6 | -7070.56 | 14261.68 | 14172.64 | 0.70 | -7045.18 | 14295.11 | 14140.29 | 0.70 | -7038.98 | 14410.61 | 14134.38 | 0.70 |
| Run 7 | -7070.47 | 14249.60 | 14166.71 | 0.70 | -7044.35 | 14296.90 | 14132.25 | 0.70 | -7037.22 | 14400.26 | 14142.53 | 0.70 |
| Run 8 | -7070.50 | 14245.17 | 14162.72 | 0.70 | -7043.33 | 14293.56 | 14130.88 | 0.70 | -7038.58 | 14392.24 | 14133.84 | 0.70 |

| Dataset II | Config 1 | | | | Config 2 | | | | Config 3 | | | |
|---|---|---|---|---|---|---|---|---|---|---|---|---|
| | EL | iBIC | iAIC | P(choice) | EL | iBIC | iAIC | P(choice) | EL | iBIC | iAIC | P(choice) |
| Run 1 | -10850.30 | 21804.32 | 21724.30 | 0.69 | -10800.67 | 21661.63 | 21643.87 | 0.69 | -10793.26 | 21930.43 | 21658.34 | 0.69 |
| Run 2 | -10852.39 | 21820.21 | 21740.18 | 0.69 | -10803.77 | 21673.18 | 21648.60 | 0.69 | -10804.72 | 21952.47 | 21680.38 | 0.69 |
| Run 3 | -10849.63 | 21799.84 | 21719.82 | 0.69 | -10798.54 | 21655.19 | 21663.95 | 0.69 | -10802.51 | 21943.64 | 21671.56 | 0.69 |
| Run 4 | -10852.65 | 21822.64 | 21742.62 | 0.68 | -10815.22 | 21689.67 | 21651.77 | 0.69 | -10798.84 | 21943.40 | 21671.32 | 0.69 |
| Run 5 | -10850.86 | 21807.71 | 21727.69 | 0.69 | -10820.94 | 21711.88 | 21649.12 | 0.69 | -10795.57 | 21939.04 | 21666.95 | 0.69 |
| Run 6 | -10853.94 | 21826.95 | 21746.93 | 0.68 | -10798.52 | 21657.83 | 21647.86 | 0.69 | -10811.31 | 21953.16 | 21681.08 | 0.69 |
| Run 7 | -10852.90 | 21816.69 | 21736.66 | 0.68 | -10798.27 | 21657.23 | 21657.37 | 0.69 | -10796.17 | 21945.84 | 21673.76 | 0.69 |
| Run 8 | -10855.17 | 21824.78 | 21744.75 | 0.68 | -10797.84 | 21650.46 | 21644.90 | 0.69 | -10800.32 | 21948.69 | 21676.61 | 0.69 |
| **Joint** | Config 1 | | | | Config 2 | | | | Config 3 | | | |
| | EL | iBIC | iAIC | P(choice) | EL | iBIC | iAIC | P(choice) | EL | iBIC | iAIC | P(choice) |
| Run 1 | -17967.33 | 36068.25 | 35983.05 | 0.69 | -17912.39 | 36061.78 | 35874.32 | 0.69 | -17895.40 | 36139.96 | 35850.25 | 0.69 |
| Run 2 | -17966.93 | 36039.00 | 35953.79 | 0.69 | -17903.43 | 36030.57 | 35843.11 | 0.69 | -17894.83 | 36138.55 | 35848.84 | 0.69 |
| Run 3 | -17965.01 | 36058.40 | 35973.19 | 0.69 | -17917.57 | 36077.21 | 35889.75 | 0.69 | -17894.20 | 36131.06 | 35841.35 | 0.69 |
| Run 4 | -17969.78 | 36075.07 | 35989.86 | 0.69 | -17912.22 | 36059.52 | 35872.06 | 0.69 | -17891.83 | 36120.43 | 35830.72 | 0.69 |
| Run 5 | -17965.26 | 36040.45 | 35955.25 | 0.69 | -17903.09 | 36013.61 | 35826.15 | 0.69 | -17898.63 | 36132.23 | 35842.53 | 0.69 |
| Run 6 | -17967.00 | 36064.37 | 35979.16 | 0.69 | -17909.19 | 36045.12 | 35857.66 | 0.69 | -17896.35 | 36144.60 | 35854.89 | 0.69 |
| Run 7 | -17966.09 | 36065.16 | 35979.95 | 0.69 | -17900.21 | 36048.40 | 35860.95 | 0.69 | -17896.23 | 36138.52 | 35848.81 | 0.69 |
| Run 8 | -17964.04 | 36058.19 | 35972.98 | 0.69 | -17916.13 | 36058.04 | 35870.58 | 0.69 | -17898.74 | 36149.77 | 35860.06 | 0.69 |

*Table 4. Group parameter estimates for configuration 2 across runs for each dataset. We see high stability in the estimated group posteriors. iBIC: integrated Bayesian information criterion, iAIC: integrated Akaike information criterion, EL: evidence lower bound, SE: standard error across runs*

| **Dataset I** | Group 1 | | | | | Group 2 | | | | |
|---|---|---|---|---|---|---|---|---|---|---|
| | lr | beta | GE | NGA | Q-decay | lr | beta | GE | NGA | Q-decay |
| Run 1 | 0.45 | 4.13 | 0.08 | -0.02 | 0.11 | 0.09 | 9.48 | 0.05 | -0.01 | 0.04 |
| Run 2 | 0.46 | 3.99 | 0.09 | -0.03 | 0.11 | 0.10 | 9.08 | 0.05 | -0.01 | 0.04 |
| Run 3 | 0.44 | 4.19 | 0.08 | -0.02 | 0.11 | 0.09 | 9.22 | 0.05 | -0.01 | 0.04 |
| Run 4 | 0.47 | 3.90 | 0.09 | -0.03 | 0.11 | 0.10 | 8.93 | 0.05 | -0.01 | 0.04 |

| | | | | | | | | | | |
|---|---|---|---|---|---|---|---|---|---|---|
| Run 5 | 0.46 | 4.02 | 0.09 | -0.03 | 0.11 | 0.10 | 8.93 | 0.05 | -0.01 | 0.04 |
| Run 6 | 0.45 | 4.11 | 0.09 | -0.02 | 0.11 | 0.10 | 9.14 | 0.05 | -0.01 | 0.04 |
| Run 7 | 0.46 | 4.01 | 0.09 | -0.03 | 0.11 | 0.10 | 8.99 | 0.05 | -0.01 | 0.04 |
| Run 8 | 0.47 | 3.87 | 0.10 | -0.03 | 0.11 | 0.10 | 8.89 | 0.05 | -0.01 | 0.04 |
| Mean | 0.46 | 4.03 | 0.09 | -0.03 | 0.11 | 0.10 | 9.08 | 0.05 | -0.01 | 0.04 |
| SE | 4.10E-03 | 3.90E-02 | 1.55E-03 | 7.06E-04 | 6.92E-04 | 1.18E-03 | 7.02E-02 | 2.58E-04 | 1.64E-04 | 2.12E-04 |
| **Dataset II** | Group 1 | | | | | Group 2 | | | | |
| | lr | beta | GE | NGA | Q-decay | lr | beta | GE | NGA | Q-decay |
| Run 1 | 0.46 | 5.56 | 0.06 | -0.07 | 0.14 | 0.16 | 10.00 | 0.00 | -0.02 | 0.05 |
| Run 2 | 0.42 | 6.19 | 0.05 | -0.07 | 0.15 | 0.19 | 8.94 | 0.01 | -0.01 | 0.05 |
| Run 3 | 0.53 | 5.03 | 0.07 | -0.08 | 0.15 | 0.19 | 9.09 | 0.00 | -0.02 | 0.06 |
| Run 4 | 0.40 | 7.19 | 0.05 | -0.06 | 0.17 | 0.18 | 9.14 | 0.00 | -0.02 | 0.06 |
| Run 5 | 1.00 | 4.21 | 0.11 | -0.09 | 0.21 | 0.21 | 9.11 | 0.01 | -0.04 | 0.09 |
| Run 6 | 0.52 | 4.82 | 0.07 | -0.09 | 0.14 | 0.18 | 9.49 | 0.00 | -0.02 | 0.06 |
| Run 7 | 0.52 | 4.81 | 0.07 | -0.09 | 0.14 | 0.18 | 9.44 | 0.00 | -0.02 | 0.06 |
| Run 8 | 0.52 | 4.78 | 0.07 | -0.09 | 0.14 | 0.19 | 9.08 | 0.00 | -0.02 | 0.06 |
| Mean | 0.55 | 5.32 | 0.07 | -0.08 | 0.15 | 0.19 | 8.79 | 0.01 | -0.02 | 0.05 |
| SE | 1.78E-01 | 8.97E-01 | 1.81E-02 | 9.70E-03 | 2.43E-02 | 4.20E-03 | 1.31E-01 | 3.48E-04 | 1.78E-03 | 2.09E-03 |
| **JOINT** | Group 1 | | | | | Group 2 | | | | |
| | lr | beta | GE | NGA | Q-decay | lr | beta | GE | NGA | Q-decay |
| Run 1 | 0.354 | 6.975 | 0.042 | -0.050 | 0.143 | 0.122 | 9.551 | 0.024 | -0.009 | 0.040 |
| Run 2 | 0.380 | 6.380 | 0.052 | -0.057 | 0.138 | 0.150 | 9.119 | 0.016 | -0.006 | 0.041 |
| Run 3 | 0.341 | 7.216 | 0.041 | -0.048 | 0.144 | 0.122 | 9.556 | 0.024 | -0.008 | 0.039 |
| Run 4 | 0.359 | 6.845 | 0.043 | -0.051 | 0.143 | 0.123 | 9.680 | 0.023 | -0.010 | 0.042 |
| Run 5 | 0.395 | 6.426 | 0.049 | -0.056 | 0.145 | 0.147 | 8.680 | 0.024 | -0.011 | 0.042 |
| Run 6 | 0.367 | 6.804 | 0.044 | -0.052 | 0.142 | 0.130 | 9.134 | 0.024 | -0.009 | 0.039 |
| Run 7 | 0.409 | 6.282 | 0.050 | -0.056 | 0.146 | 0.142 | 8.861 | 0.024 | -0.014 | 0.044 |
| Run 8 | 0.349 | 7.056 | 0.041 | -0.049 | 0.144 | 0.123 | 9.577 | 0.024 | -0.010 | 0.041 |
| Mean | 0.369 | 6.748 | 0.045 | -0.052 | 0.143 | 0.132 | 9.270 | 0.023 | -0.010 | 0.041 |
| SE | 7.41E-03 | 1.08E-01 | 1.37E-03 | 1.04E-03 | 7.28E-04 | 3.74E-03 | 1.17E-01 | 9.08E-04 | 6.73E-04 | 5.22E-04 |

##### 1.3.2 Recoverability of single fitted datasets

*Table 5 ICC- Parameter recoverability for each identified cluster. Synthetic data were generated from the hierarchically fitted group posteriors (single simulation run) and refitted with the same model. ICC values are reported for each cluster, as identified for single dataset fits. std: standard deviation*

| | Dataset I (Single Fit) | | | | Dataset II (Single Fit) | | | |
|---|---|---|---|---|---|---|---|---|
| | Cluster 1 | | Cluster 2 | | Cluster 1 | | Cluster 2 | |
| | mean | std | mean | std | mean | std | mean | std |
| Learning Rate | 0.49 | 0.16 | 0.80 | 0.04 | 0.04 | 0.02 | 0.49 | 0.07 |
| Inverse Temp | 0.14 | 0.10 | -0.04 | 0.12 | 0.01 | 0.01 | 0.01 | 0.03 |
| Escape Bias | 0.62 | 0.10 | 0.87 | 0.02 | 0.45 | 0.05 | 0.76 | 0.03 |
| Avoid Bias | 0.63 | 0.14 | 0.88 | 0.03 | 0.38 | 0.08 | 0.78 | 0.02 |
| Q-decay | 0.89 | 0.04 | 0.80 | 0.07 | 0.11 | 0.04 | 0.62 | 0.05 |

##### 1.3.3 Parameter Correlation tables

To further investigate the validity of the model parameterization, we computed the posterior correlation of the parameters in the hierarchically fitted models and averaged across participants. Correlations are small, except for notable correlations between learning rate and Q-Decay in both datasets. Both parameters high recoverability and model selection clearly justifying the inclusion of this parameter, show that both capture unique variances in the assessed data.

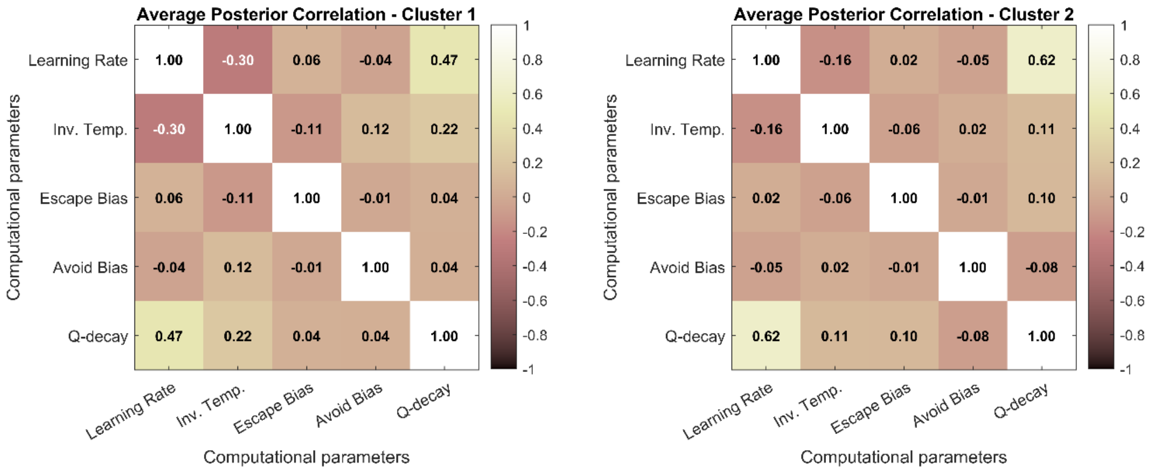

*Figure 1-3 Posterior parameter correlations, averaged over 8 runs of K=2 configuration, fitted on datasets I and II jointly.*

##### 1.3.4 Comparison between the clusters found in Joint and Single Dataset Modelling

To assess the correspondence between joint and single-dataset clustering solutions, we compared participant assignments after cluster label alignment. In Dataset I, cluster assignments differed for 6 subjects (12%). In Dataset II, assignments differed for 29 subjects (14.7%). However, 14 of these individuals were not robustly assigned in either the joint clustering (n = 5) or the single-dataset clustering (n = 9), indicating ambiguous cluster membership. The remaining 15 subjects were confidently assigned to different clusters across approaches. After alignment, we see an overall high recovery of cluster assignment for both datasets in the joint fit. The highest confusion rate is found in Dataset II, cluster 2, where only 63% of participants are reassigned to the joint cluster 2 (Fig 1-4).

Overall, the joint analysis showed high recovery of cluster assignments across datasets, indicating that the behavioural regimes identified in the single-dataset analyses largely correspond to shared computational profiles. The highest confusion occurred for Dataset II Cluster 2, where only 63% of participants were reassigned to the same cluster in the joint solution (Fig. 1-4).

To assess parameter consistency between joint and single dataset fits further, we computed intraclass correlation coefficients (ICC) for individuals assigned to the same cluster across analyses (Table 5). ICC values indicated strong agreement in the parametrization of the clusters between joint and single fits. As in the previous analyses, the inverse temperature parameter (β) showed weaker recoverability relative to the other parameters.

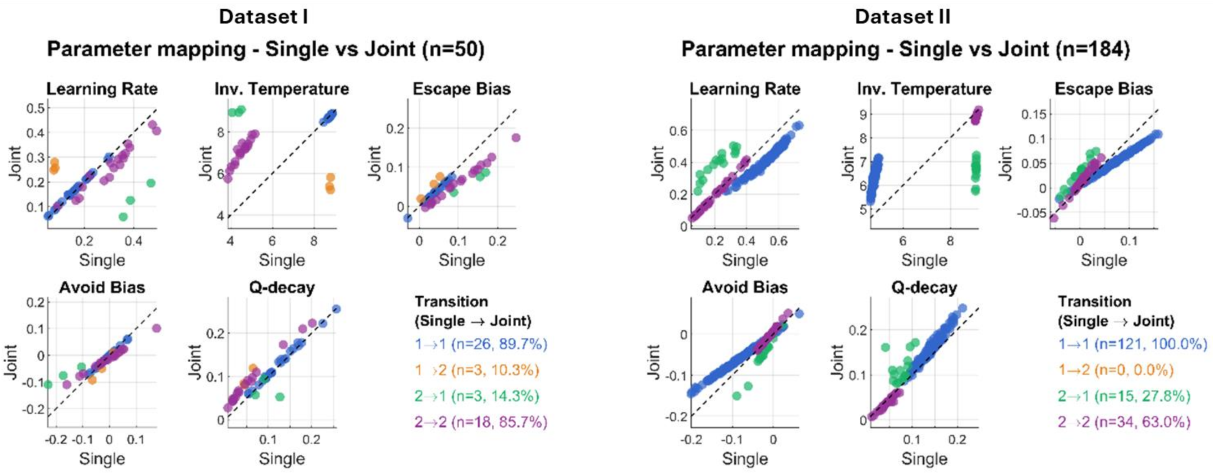

*Figure 1-4 Jointly fitted model for Dataset I and II. For each original dataset, we show how the original clusters map to the joint clusters. Each panel shows the correspondence between parameter estimates, with data points colour-coded according to cluster assignment transitions. Subjects not assigned to a cluster either in single or join fitting are not shown.*

*Table 6 ICC- values for subjects reassigned in the same cluster in single vs joint fits*

| | Dataset I | | Dataset II | |
|---|---|---|---|---|
| Single assignment – Joint Assignment | 1-1 | 2-2 | 1-1 | 2-2 |
| Learning Rate | 0.99 | 0.87 | 0.55 | 0.99 |
| Inverse Temperature | 0.97 | 0.06 | 0.02 | 0.1 |
| Escape Bias | 0.98 | 0.77 | 0.8 | 0.87 |
| Avoid Bias | 0.99 | 0.89 | 0.81 | 0.95 |
| Q-Decay | 1.00 | 0.88 | 0.84 | 0.88 |

Together with the single-dataset analyses, these results indicate that the fast/adaptive and slow/perseverative regimes generalize across datasets and task variants.

### 1.4 Cluster Classification

#### 1.4.1 Clinical Measures across datasets

We assessed the distribution of clinical variables within each cluster (per dataset) and used two-sided t-tests with Bonferroni correction to identify any significant differences between the clusters. We found no significant differences between the investigated clusters in either dataset.

*Table 7. Dataset I – Clinical Statistics and Comparison for cluster 1 and cluster 2 (as identified in joint fitting)*

| | Median_Cluster1 | Q25_Cluster1 | Q75_Cluster1 | Median_Cluster2 | Q25_Cluster2 | Q75_Cluster2 | p_bonferroni | Cohens_d |
|---|---|---|---|---|---|---|---|---|
| SI | 4 | 2.5 | 5 | 3 | 1.75 | 5 | 1 | 0.270 |
| SI_sev | 16 | 13 | 17 | 15 | 12.5 | 16.5 | 1 | 0.297 |
| SI_pm | 0 | 0 | 1 | 0 | 0 | 1 | 1 | 0.181 |
| SB | 0 | 0 | 1 | 0 | 0 | 1 | 1 | 0.107 |
| attempt_no | 0 | 0 | 1 | 0 | 0 | 1 | 1 | -0.380 |
| BIS11 | 67 | 61.5 | 75.25 | 65 | 52.5 | 74.5 | 1 | 0.324 |
| BDI | 19 | 12.5 | 31.5 | 18 | 10.5 | 35.5 | 1 | -0.101 |
| BPAQ | 41 | 29.25 | 59.5 | 53 | 30 | 65.75 | 1 | -0.152 |
| ACE | 11 | 7 | 14.25 | 10 | 5.75 | 13.75 | 1 | 0.108 |
| GAD7 | 9 | 5 | 13.25 | 9 | 4 | 13.25 | 1 | 0.032 |
| BAI | 17 | 8 | 22.25 | 8 | 6.75 | 18 | 1 | 0.191 |
| STHS-S | 25 | 24 | 26 | 26 | 23.75 | 27 | 1 | -0.086 |
| STHS-T | 33 | 32 | 35 | 32 | 30.75 | 33.25 | 1 | 0.368 |

*Table 8. Dataset II – Clinical Statistics and Comparison for cluster 1 and cluster 2 (as identified in joint fitting)*

| | Median_Cluster1 | Q25_Cluster1 | Q75_Cluster1 | Median_Cluster2 | Q25_Cluster2 | Q75_Cluster2 | p_bonferroni | Cohens_d |
|---|---|---|---|---|---|---|---|---|
| hitop_ext_anankastia | 11.4 | 9.4 | 13.2 | 10.8 | 9.6 | 12.2 | 1 | 0.10 |
| hitop_ext_antagonism | 7.88 | 6.75 | 9.38 | 8 | 7.25 | 9.5 | 1 | -0.10 |
| hitop_ext_antisocial_disinhibition | 8 | 6.775 | 9.7 | 7.55 | 6.6 | 9.1 | 1 | 0.15 |
| hitop_int_distress | 8 | 6.15 | 9.58 | 7.75 | 6.25 | 9.17 | 1 | 0.12 |
| hitop_int_fear | 7.77 | 6.29 | 9 | 7.69 | 6.69 | 8.77 | 1 | -0.02 |
| inq_grand_sum | 39 | 27 | 59.25 | 40.5 | 23 | 51 | 1 | 0.12 |
| hitop_suicidality | 5 | 4 | 7 | 5 | 4 | 6 | 1 | 0.18 |
| gcsq_perceived_capability | 3 | 1.75 | 4 | 3 | 1 | 4 | 1 | 0.01 |
| gcsq_pain_tolerance | 12 | 9 | 15 | 13.5 | 10 | 14 | 1 | -0.12 |

| | | | | | | | | |
|---|---|---|---|---|---|---|---|---|
| gcsq_fearlesness_of_death | 14 | 7.75 | 21 | 13 | 7 | 21 | 1 | 0.10 |

##### 1.4.2 Computational Parameters across datasets

Two further investigate how the two datasets are modelled in the joint fit, we assess if there are statistically significant differences in the parameters for participants from the two datasets. We did find significant differences in the Avoid bias, which while statistically significant is a small absolute difference, and in the learning rate. The learning rate is higher in the more volatile task, as participants have fewer trials to learn and relearn the environment in the task deployed for dataset II.

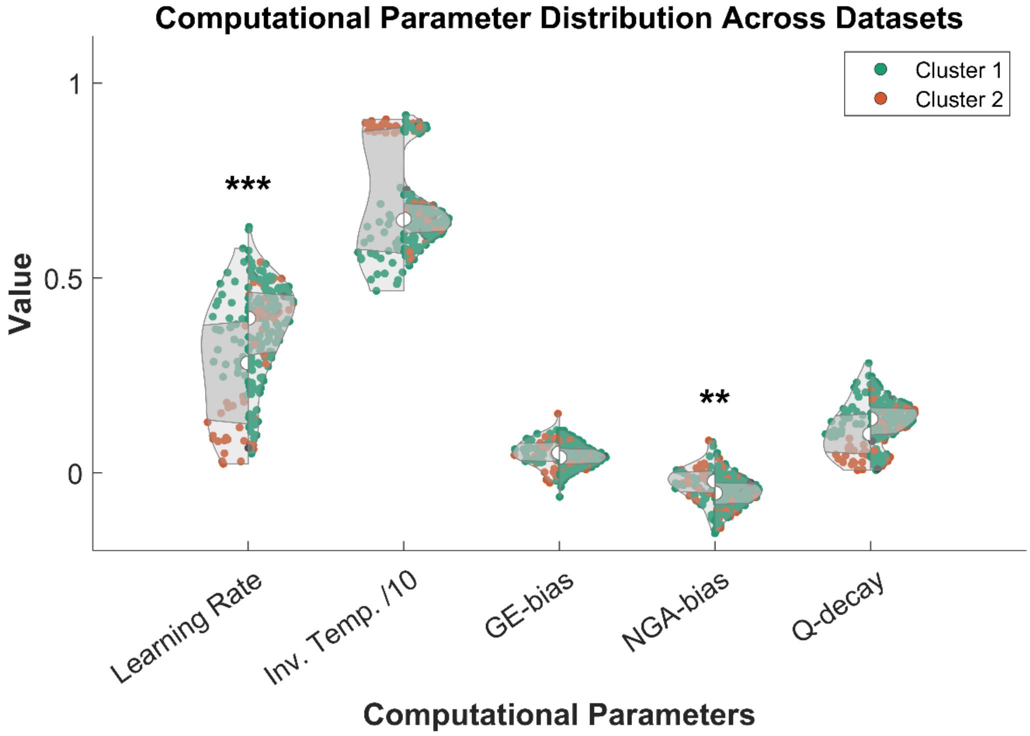

*Figure 1-5 Computational parameters differ across datasets. Fitted parameters are shown for dataset I (left) and dataset II (right). Scattered markers are colour marked based on the cluster assignment of the specific subject. After Bonferroni correction, two differences remain statistically significant; Learning Rate (P_bonf = 1.63e-05, d = -0.763) and NGA-bias (P_bonf = 4.79e-04, d = 0.635).*

##### 1.4.3 Full Behavioural Traces

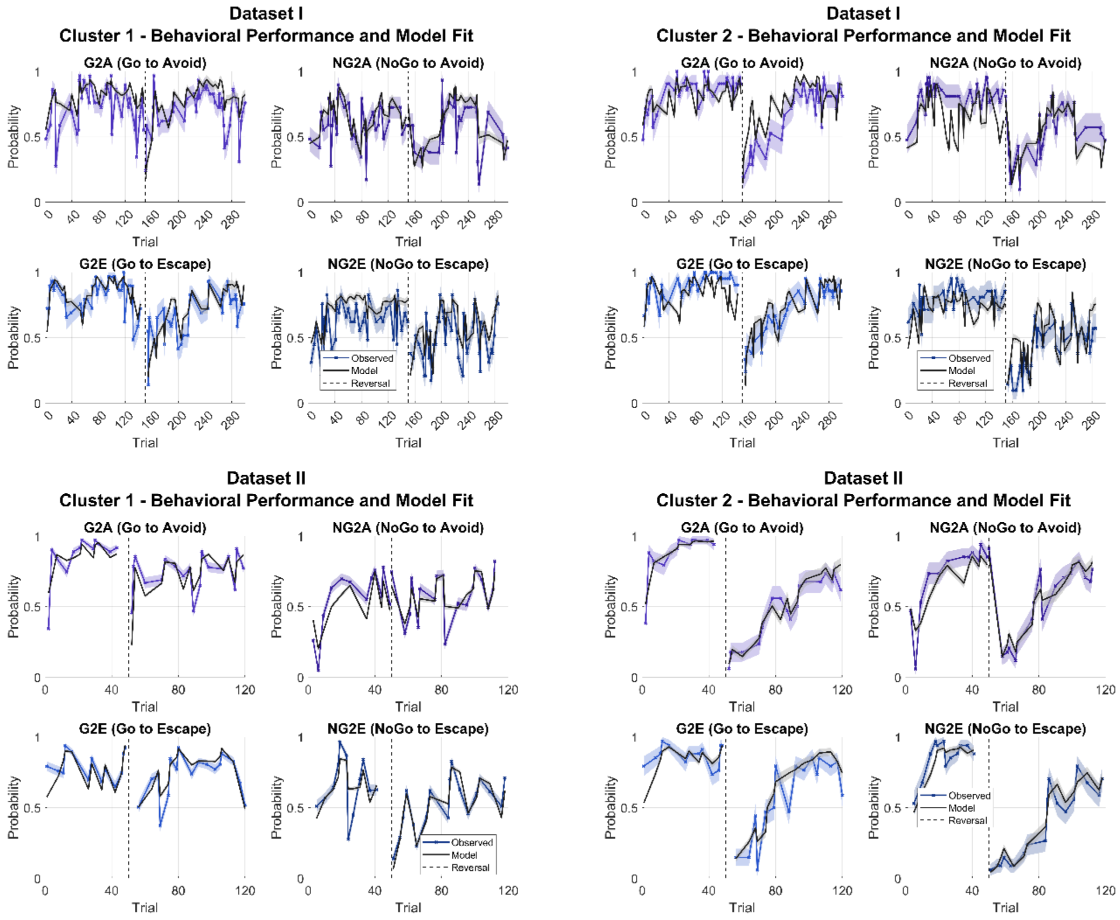

*Figure 1-6* ***Full behavioural traces across clusters and dataset.***

#### 1.4.4 CCA Details

##### *1.4.4.1 Weights*

This section contains the details of the Canonical Variates of the CCA analyses at population and cluster levels. Weights for each variable (computational parameters, clinical and demographic) are detailed, as well as average weights for the clinical subfactors reported for dataset II.

###### 1.4.4.1.1 Dataset I

*Table 9 Dataset I - Population-level CCA, weights for the first canonical variate (CV1), including computational parameters and clinical variates. Sorted in descending order of the associated weights.*

| Variables | Weights – CV1 |
|---|---|
| **Avoid Bias** | 0.332 |
| **Q-decay** | -0.009 |
| **Learning Rate** | -0.125 |
| **Inv. Temp.** | -0.401 |
| **Escape Bias** | -0.778 |
| **sex** | 0.080 |
| **age** | 0.028 |
| **C_SSRS_ideation** | -0.021 |
| **ACE** | -0.035 |
| **C_SSRS_ideation_sev** | -0.066 |
| **C_SSRS_attempt_no** | -0.215 |
| **BIS11** | -0.446 |
| **BPAQ** | -0.476 |
| **C_SSRS_behav** | -0.521 |
| **STHS-trait** | -0.554 |
| **STHS-state** | -0.578 |
| **C_SSRS_ideation_sev_pm** | -0.698 |
| **GAD7** | -0.736 |
| **BAI-T** | -0.761 |
| **BDI** | -0.904 |

*Table 10 Dataset I – Cluster 1 (fast/adaptive) CCA, weights for the first canonical variate(CV1), including computational parameters and clinical variates. Sorted in descending order of the associated weights.*

| Variable | Weight - CV1 |
|---|---|
| **Q-decay** | 0.340 |
| **Avoid Bias** | 0.154 |
| **Inv. Temp.** | -0.305 |
| **Learning Rate** | -0.437 |
| **Escape Bias** | -0.680 |
| **Sex - Assigned** | 0.348 |
| **Age** | 0.073 |
| **ACE** | 0.068 |
| **C_SSRS_attempt_no** | -0.244 |
| **C_SSRS_ideation** | -0.313 |
| **C_SSRS_ideation_sev** | -0.342 |
| **STHS-state** | -0.400 |
| **STHS-trait** | -0.449 |
| **BIS11** | -0.525 |
| **C_SSRS_behav** | -0.657 |
| **BPAQ** | -0.658 |
| **C_SSRS_ideation_sev_pm** | -0.673 |
| **BAI-T** | -0.749 |
| **GAD7** | -0.788 |
| **BDI** | -0.868 |

*Table 11 Dataset I – Cluster 2 (slow/perseverative) CCA, weights for the first canonical variate(CV1), including computational parameters and clinical variates. Sorted in descending order of the associated weights.*

| Variable | Weight - CV1 |
|---|---|
| **Learning Rate** | 0.665 |
| **Avoid Bias** | 0.587 |
| **Escape Bias** | -0.026 |
| **Inv. Temp.** | -0.301 |
| **Q-decay** | -0.652 |
| **C_SSRS_ideation** | 0.504 |
| **C_SSRS_ideation_sev** | 0.443 |
| **Sex- assigned** | 0.315 |
| **ACE** | 0.072 |
| **C_SSRS_attempt_no** | -0.152 |
| **BPAQ** | -0.171 |
| **C_SSRS_behav** | -0.216 |
| **BIS11** | -0.339 |
| **Age** | -0.388 |
| **GAD7** | -0.763 |
| **STHS-trait** | -0.773 |
| **C_SSRS_ideation_sev_pm** | -0.848 |
| **STHS-state** | -0.849 |
| **BAI-T** | -0.907 |
| **BDI** | -0.950 |

###### 1.4.4.1.2 Dataset II

*Table 12 Dataset II – population-level CCA, weights for the first canonical variate. Clinical variables are averaged in subcategories for clinical variables. Weights on the individual clinical variable level are displayed in a separate table 13 . Sorted in descending order.*

| Variable | Section | Weight-CV1 |
|---|---|---|
| **Avoid Bias** | PARAMETER | 0.332 |
| **Q-decay** | PARAMETER | -0.009 |
| **Learning Rate** | PARAMETER | -0.125 |
| **Inv. Temp.** | PARAMETER | -0.401 |
| **Escape Bias** | PARAMETER | -0.778 |
| **INQ** | SUICIDALITY | 0.801 |
| **GCSQ Fearless. Death** | SUICIDALITY | 0.347 |
| **Antisocial/Disinhibition** | EXTERNALIZING | 0.233 |
| **Antagonism** | EXTERNALIZING | 0.12 |
| **Distress** | INTERNALIZING | 0.115 |
| **Fear** | INTERNALIZING | 0.108 |
| **Active Mental Health Treatment** | DEMOGRAPHICS | 0.033 |
| **Anankastia** | EXTERNALIZING | -0.1 |
| **GCSQ Pain Tol.** | SUICIDALITY | -0.159 |
| **Sex Assigned** | DEMOGRAPHICS | -0.304 |
| **Age** | DEMOGRAPHICS | -0.439 |
| **GCSQ Perc. Cap.** | SUICIDALITY | -0.597 |

*Table 13 Dataset II – population-level CCA: weights for individual clinical factors in descending order.*

| Clinical/demographic variable | Subfactor | Weight – CV1 |
|---|---|---|
| **INQ** | Suicidality - explicit | 0.801 |
| **Nssi** | Internalizing - Distress | 0.722 |
| **Distress Dysphoria Anhedonia** | Internalizing - Distress | 0.61 |
| **Distress Dysphoria Depressed Mood** | Internalizing - Distress | 0.61 |
| **Antisocial Behaviour** | Externalizing - Antisocial/Disinhibition | 0.556 |
| **Specific Phobia Blood Injection** | Internalizing - Fear | 0.497 |
| **Gambling** | Externalizing - Antisocial/Disinhibition | 0.488 |
| **Distress Dysphoria Shame Guilt** | Internalizing - Distress | 0.414 |
| **Callousness** | Externalizing - Antagonism | 0.404 |
| **Gaming** | Externalizing - Antisocial/Disinhibition | 0.388 |
| **GCSQ Fearless. Death** | Suicidality - explicit | 0.347 |
| **Dishonesty Deceitfulness** | Externalizing - Antagonism | 0.323 |
| **Excoriation** | Internalizing - Fear | 0.312 |
| **Entitlement** | Externalizing - Antagonism | 0.304 |
| **Risk Taking** | Externalizing - Antisocial/Disinhibition | 0.268 |
| **Dishonesty Manipulativeness** | Externalizing - Antagonism | 0.254 |
| **Trichotillomania** | Internalizing - Fear | 0.231 |

| | | |
|---|---|---|
| **Disorganization** | Externalizing - Antisocial/Disinhibition | 0.216 |
| **Distress Dysphoria Lassitude** | Internalizing - Distress | 0.205 |
| **Nightmares** | Internalizing - Distress | 0.194 |
| **Workaholism** | Externalizing - Anankastia | 0.186 |
| **Counting** | Internalizing - Fear | 0.186 |
| **Panic** | Internalizing - Fear | 0.162 |
| **Trauma Reactions** | Internalizing - Fear | 0.154 |
| **Nonplanfulness** | Externalizing - Antisocial/Disinhibition | 0.146 |
| **Cleaning** | Internalizing - Fear | 0.137 |
| **Oppositionality** | Externalizing - Antisocial/Disinhibition | 0.111 |
| **Social Aggression** | Externalizing - Antagonism | 0.109 |
| **Nonpersistence** | Externalizing - Antisocial/Disinhibition | 0.101 |
| **Exhibitionism** | Externalizing - Antagonism | 0.082 |
| **Restlessness** | Externalizing - Antisocial/Disinhibition | 0.062 |
| **Specific Phobia Animal Insect** | Internalizing - Fear | 0.062 |
| **Perfectionism** | Externalizing - Anankastia | 0.03 |
| **Emotionality Affective Lability** | Internalizing - Distress | 0.001 |
| **Problematic Shopping** | Externalizing - Antisocial/Disinhibition | 0 |
| **Agoraphobia** | Internalizing - Fear | -0.007 |
| **Cognitive Problems** | Internalizing - Distress | -0.025 |
| **Checking** | Internalizing - Fear | -0.032 |
| **Hoarding** | Internalizing - Fear | -0.039 |
| **Rigidity** | Externalizing - Anankastia | -0.054 |
| **Specific Phobia Situational** | Internalizing - Fear | -0.064 |
| **GCSQ Pain Tol.** | Suicidality - explicit | -0.159 |
| **Domineering** | Externalizing - Antagonism | -0.175 |
| **Distress Dysphoria Anxious Worry** | Internalizing - Distress | -0.181 |
| **Insomnia** | Internalizing - Distress | -0.186 |
| **Hypervigilance** | Internalizing - Fear | -0.199 |
| **Hyperdeliberation** | Externalizing - Anankastia | -0.263 |
| **Grandiosity** | Externalizing - Antagonism | -0.344 |
| **Risk Aversion** | Externalizing - Anankastia | -0.397 |
| **Emotionality Angry Hostility** | Internalizing - Distress | -0.454 |
| **Emotionality Irritability** | Internalizing - Distress | -0.528 |
| **GCSQ Perc. Cap.** | Suicidality - explicit | -0.597 |

*Table 14 Dataset II – Cluster 1 (fast/adaptive) CCA, weights for the first canonical variate. Clinical variables are averaged in subcategories for clinical variables. Weights on the individual clinical variable level are displayed in a separate table 15 . Sorted in descending order*

| Variable | Weight - CV1 | |
|---|---|---|
| **Q-decay** | PARAMETER | 0.340 |
| **Avoid Bias** | PARAMETER | 0.154 |
| **Inv. Temp.** | PARAMETER | -0.305 |
| **Learning Rate** | PARAMETER | -0.437 |
| **Escape Bias** | PARAMETER | -0.680 |
| **INQ** | SUICIDALITY | 0.516 |
| **GCSQ Fearless. Death** | SUICIDALITY | 0.380 |
| **Active Mental Health Treatment** | DEMOGRAPHICS | 0.274 |
| **Antagonism** | EXTERNALIZING | 0.111 |
| **Age** | DEMOGRAPHICS | -0.002 |
| **Antisocial/Disinhibition** | EXTERNALIZING | -0.066 |
| **Distress** | INTERNALIZING | -0.077 |
| **GCSQ Pain Tol.** | SUICIDALITY | -0.133 |
| **Fear** | INTERNALIZING | -0.200 |
| **GCSQ Perc. Cap.** | SUICIDALITY | -0.216 |
| **Sex Assigned** | DEMOGRAPHICS | -0.275 |
| **Anankastia** | EXTERNALIZING | -0.282 |

*Table 15 Dataset II – Cluster 1 (fast/adaptive) CCA, weights individual clinical factors for the first canonical variate in descending order.*

| Clinical/demographic variable | Subfactor | Weight – CV1 |
|---|---|---|
| **Nssi** | Internalizing - Distress | 0.565 |
| **Antisocial Behaviour** | Externalizing - Antisocial/Disinhibition | 0.554 |
| **Callousness** | Externalizing - Antagonism | 0.52 |
| **INQ** | Suicidality - explicit | 0.516 |
| **GCSQ Fearless. Death** | Suicidality - explicit | 0.38 |
| **Social Aggression** | Externalizing - Antagonism | 0.362 |
| **Distress Dysphoria Depressed Mood** | Internalizing - Distress | 0.355 |
| **Distress Dysphoria Anhedonia** | Internalizing - Distress | 0.289 |
| **Gambling** | Externalizing - Antisocial/Disinhibition | 0.263 |
| **Risk Taking** | Externalizing - Antisocial/Disinhibition | 0.247 |
| **Entitlement** | Externalizing - Antagonism | 0.189 |
| **Dishonesty Manipulativeness** | Externalizing - Antagonism | 0.157 |
| **Oppositionality** | Externalizing - Antisocial/Disinhibition | 0.153 |
| **Trichotillomania** | Internalizing - Fear | 0.136 |
| **Exhibitionism** | Externalizing - Antagonism | 0.075 |
| **Gaming** | Externalizing - Antisocial/Disinhibition | 0.014 |
| **Specific Phobia Blood Injection** | Internalizing - Fear | 0.007 |

| | | |
|---|---|---|
| **Distress Dysphoria Shame Guilt** | Internalizing - Distress | 0.001 |
| **Cleaning** | Internalizing - Fear | -0.007 |
| **Emotionality Affective Lability** | Internalizing - Distress | -0.038 |
| **Counting** | Internalizing - Fear | -0.043 |
| **Emotionality Angry Hostility** | Internalizing - Distress | -0.05 |
| **Domineering** | Externalizing - Antagonism | -0.054 |
| **Trauma Reactions** | Internalizing - Fear | -0.086 |
| **Agoraphobia** | Internalizing - Fear | -0.088 |
| **Dishonesty Deceitfulness** | Externalizing - Antagonism | -0.098 |
| **Nonplanfulness** | Externalizing - Antisocial/Disinhibition | -0.126 |
| **GCSQ Pain Tol.** | Suicidality - explicit | -0.133 |
| **Workaholism** | Externalizing - Anankastia | -0.15 |
| **Rigidity** | Externalizing - Anankastia | -0.151 |
| **Excoriation** | Internalizing - Fear | -0.168 |
| **Distress Dysphoria Lassitude** | Internalizing - Distress | -0.19 |
| **Nightmares** | Internalizing - Distress | -0.192 |
| **Panic** | Internalizing - Fear | -0.199 |
| **Specific Phobia Animal Insect** | Internalizing - Fear | -0.213 |
| **GCSQ Perc. Cap.** | Suicidality - explicit | -0.216 |
| **Restlessness** | Externalizing - Antisocial/Disinhibition | -0.239 |
| **Grandiosity** | Externalizing - Antagonism | -0.262 |
| **Emotionality Irritability** | Internalizing - Distress | -0.274 |
| **Hyperdeliberation** | Externalizing - Anankastia | -0.286 |
| **Specific Phobia Situational** | Internalizing - Fear | -0.322 |
| **Distress Dysphoria Anxious Worry** | Internalizing - Distress | -0.358 |
| **Risk Aversion** | Externalizing - Anankastia | -0.403 |
| **Insomnia** | Internalizing - Distress | -0.408 |
| **Perfectionism** | Externalizing - Anankastia | -0.422 |
| **Hypervigilance** | Internalizing - Fear | -0.431 |
| **Problematic Shopping** | Externalizing - Antisocial/Disinhibition | -0.435 |
| **Disorganization** | Externalizing - Antisocial/Disinhibition | -0.512 |
| **Hoarding** | Internalizing - Fear | -0.576 |
| **Nonpersistence** | Externalizing - Antisocial/Disinhibition | -0.577 |
| **Checking** | Internalizing - Fear | -0.615 |
| **Cognitive Problems** | Internalizing - Distress | -0.628 |

*Table 16 Dataset II – Cluster 2 (slow/ perseverative) CCA, weights for the first canonical variate. Clinical variables are averaged in subcategories for clinical variables. Weights on the individual clinical variable level are displayed in a separate table 17 . Sorted in descending order*

| Variable | Section | Weight-CV1 |
|---|---|---|
| **Learning Rate** | PARAMETER | 0.665 |
| **Avoid Bias** | PARAMETER | 0.587 |
| **Escape Bias** | PARAMETER | -0.026 |
| **Inv. Temp.** | PARAMETER | -0.301 |
| **Q-decay** | PARAMETER | -0.652 |
| **GCSQ Perc. Cap.** | SUICIDALITY | 0.534 |
| **GCSQ Pain Tol.** | SUICIDALITY | 0.234 |
| **Age** | DEMOGRAPHICS | 0.169 |
| **Active Mental Health Treatment** | DEMOGRAPHICS | -0.017 |
| **GCSQ Fearless. Death** | SUICIDALITY | -0.234 |
| **Anankastia** | EXTERNALIZING | -0.308 |
| **Sex Assigned** | DEMOGRAPHICS | -0.33 |
| **Antagonism** | EXTERNALIZING | -0.439 |
| **INQ** | SUICIDALITY | -0.477 |
| **Antisocial/Disinhibition** | EXTERNALIZING | -0.544 |
| **Fear** | INTERNALIZING | -0.546 |
| **Distress** | INTERNALIZING | -0.639 |

*Table 17 Dataset II – Cluster 2 (slow/ perseverative) CCA: weights for individual clinical factors in descending order*

| Clinical/demographic variable | Subfactor | Weight – CV1 |
|---|---|---|
| **GCSQ Perc. Cap.** | Suicidality - explicit | 0.534 |
| **GCSQ Pain Tol.** | Suicidality - explicit | 0.234 |
| **Grandiosity** | Externalizing - Antagonism | 0.195 |
| **Hyperdeliberation** | Externalizing - Anankastia | -0.036 |
| **Risk Aversion** | Externalizing - Anankastia | -0.056 |
| **Trichotillomania** | Internalizing - Fear | -0.075 |
| **Domineering** | Externalizing - Antagonism | -0.191 |
| **Gambling** | Externalizing - Antisocial/Disinhibition | -0.194 |
| **GCSQ Fearless. Death** | Suicidality - explicit | -0.234 |
| **Nightmares** | Internalizing - Distress | -0.296 |
| **Antisocial Behaviour** | Externalizing - Antisocial/Disinhibition | -0.308 |
| **Workaholism** | Externalizing - Anankastia | -0.325 |
| **Callousness** | Externalizing - Antagonism | -0.355 |
| **Exhibitionism** | Externalizing - Antagonism | -0.383 |
| **Gaming** | Externalizing - Antisocial/Disinhibition | -0.395 |
| **INQ** | Suicidality - explicit | -0.477 |
| **Risk Taking** | Externalizing - Antisocial/Disinhibition | -0.483 |
| **Specific Phobia Animal Insect** | Internalizing - Fear | -0.488 |

| | | |
|---|---|---|
| **Distress Dysphoria Anhedonia** | Internalizing - Distress | -0.498 |
| **Excoriation** | Internalizing - Fear | -0.499 |
| **Perfectionism** | Externalizing - Anankastia | -0.504 |
| **Cleaning** | Internalizing - Fear | -0.525 |
| **Restlessness** | Externalizing - Antisocial/Disinhibition | -0.53 |
| **Specific Phobia Situational** | Internalizing - Fear | -0.537 |
| **Agoraphobia** | Internalizing - Fear | -0.561 |
| **Counting** | Internalizing - Fear | -0.566 |
| **Specific Phobia Blood Injection** | Internalizing - Fear | -0.574 |
| **Hypervigilance** | Internalizing - Fear | -0.596 |
| **Nssi** | Internalizing - Distress | -0.607 |
| **Social Aggression** | Externalizing - Antagonism | -0.615 |
| **Rigidity** | Externalizing - Anankastia | -0.618 |
| **Trauma Reactions** | Internalizing - Fear | -0.623 |
| **Oppositionality** | Externalizing - Antisocial/Disinhibition | -0.632 |
| **Distress Dysphoria Lassitude** | Internalizing - Distress | -0.642 |
| **Nonpersistence** | Externalizing - Antisocial/Disinhibition | -0.645 |
| **Insomnia** | Internalizing - Distress | -0.649 |
| **Cognitive Problems** | Internalizing - Distress | -0.66 |
| **Panic** | Internalizing - Fear | -0.666 |
| **Emotionality Affective Lability** | Internalizing - Distress | -0.686 |
| **Checking** | Internalizing - Fear | -0.686 |
| **Entitlement** | Externalizing - Antagonism | -0.688 |
| **Dishonesty Manipulativeness** | Externalizing - Antagonism | -0.697 |
| **Hoarding** | Internalizing - Fear | -0.699 |
| **Distress Dysphoria Depressed Mood** | Internalizing - Distress | -0.709 |
| **Emotionality Angry Hostility** | Internalizing - Distress | -0.716 |
| **Emotionality Irritability** | Internalizing - Distress | -0.731 |
| **Distress Dysphoria Anxious Worry** | Internalizing - Distress | -0.737 |
| **Distress Dysphoria Shame Guilt** | Internalizing - Distress | -0.737 |
| **Problematic Shopping** | Externalizing - Antisocial/Disinhibition | -0.741 |
| **Nonplanfulness** | Externalizing - Antisocial/Disinhibition | -0.754 |
| **Disorganization** | Externalizing - Antisocial/Disinhibition | -0.76 |
| **Dishonesty Deceitfulness** | Externalizing - Antagonism | -0.781 |

##### 1.4.4.2 Accuracy correlation

For each cluster, we examined the correlation of the clinical and parameter canonical variates with performance accuracy (total, before reversal and after reversal) with a correlation analysis (Pearson's R). After Bonferroni correction for multiple comparisons, only the canonical variate analysed for dataset I, cluster 2 retained a significant correlation of the parameter covariate with task total accuracy (r= 0.723, p=3.2e-04) as well as accuracy before reversal (p= 0.808, p=1.6e-05).

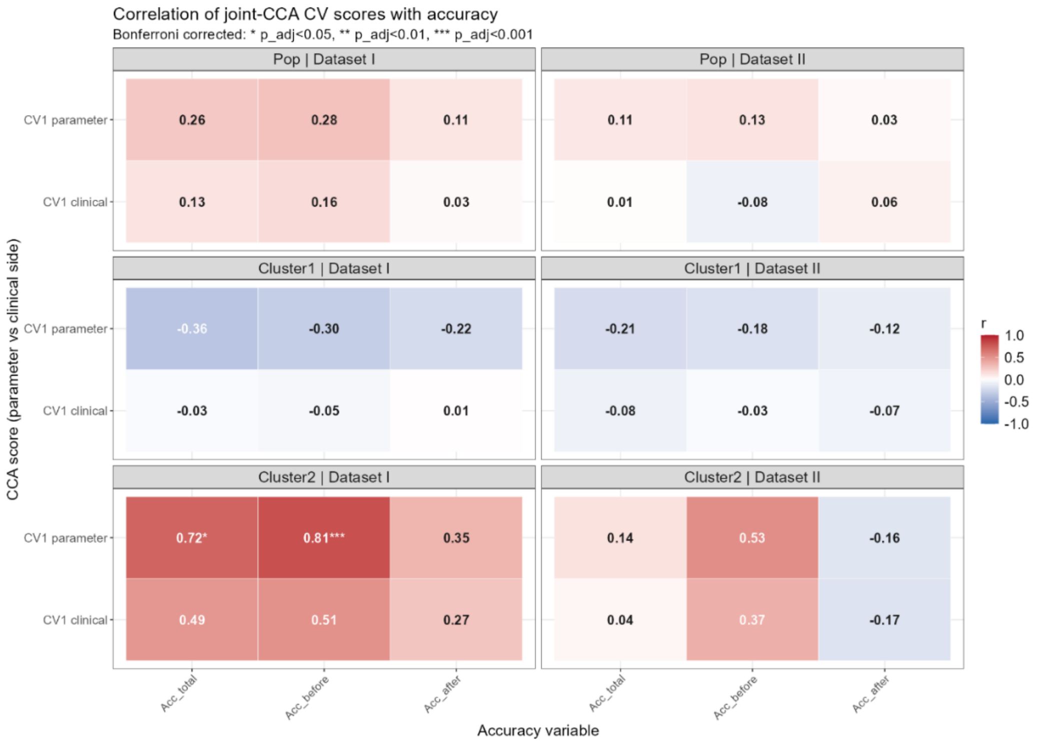

*Figure 1-7 Correlation of Canonical Covariates with Accuracy measures (total, before and after reversal).*

##### 1.4.4.3 Bootstrapping results

To assess the robustness of CCA results on each level (population, cluster 1, cluster 2), non-parametric bootstrapping with N=1000 was performed. Across iterations, the weights with which clinical variables/subfactors and parameters contributed to the recovered canonical variates was assessed and ranked against the other clinical variables/subfactors or parameters. We then assessed the frequency with which clinical variables/subfactors and parameters emerged as top contributors or within the top 20% of contributing factors as a measure of robustness.

To establish whether a factor ranked significantly higher than chance, we derived significance thresholds based on exact binomial distributions under chance performance, using a one-sided right-tail criterion with $\alpha$ = 0.05. Chance performance of the top 20% criterion was based on the number of candidate variables. For parameter canonical variates, five parameters were considered, leading to a chance performance of exactly p_null, p = .2. For dataset I, 13 clinical variables were considered, leading to $p_{\text{null,I}} = \max_{0 \le x \le 15; x \in \mathbf{Z}} \left[1 - \frac{x}{15} \le .2\right] = .222$. For dataset II, 9 clinical subfactors and suicidality variables were considered, leading to $p_{\text{null,II}} \max_{0 \le x \le 12; x \in \mathbf{Z}} [1 - \frac{x}{12} \le .2] = .187$.

This approach yields significance thresholds of 22.2% for parameter contribution in the Top 20% across datasets, 22.2% for dataset I clinical variables Top 20% and 18.7% for dataset II clinical subfactors/suicidality variables.

At the population level, bootstrapping confirms the strong weight of Escape bias on CV1, as in the fit to the full data. Similarly, for dataset I, the consistently highest ranked clinical factors confirm strong loading on depression and anxiety indicators with a secondary loading on suicidality metrics, albeit their prevalence varies in the bootstrapping results (Table 18). For dataset II, the strong weight on INQ is confirmed (Table 19). On the individual variable level, the same variables cited in the full analysis as top contributors emerge in the top 20 often when ranked at the individual clinical variable level (NSSI:51.8%, antisocial behaviour:38.1% and internalizing distress features including depressed mood:72.7%, and anhedonia:47.7%), indicating high stability.

*Table 18 Dataset I - Population bootstrapping results. Shown are the statistics across run for all computational, clinical and demographic categories. We computed the number of variables per category, the average rank across all runs, the percentage of runs in which the category scored highest among either computational or clinical/demographic category and the percentage of runs in which the category scored within the top 20 percent of categories based on the absolute weight on the first clinical covariate. Statistically significant category contributions are bolded for computational or clinical/demographic category separately.*

| Variable/Factor | N_variables | Mean_Rank | Rank1_Pct | Top20_Pct |
|---|---|---|---|---|
| **Escape Bias** | 1 | 2.05 | 49.7 | **49.7** |
| Avoid Bias | 1 | 3.06 | 17.3 | 17.3 |
| Learning Rate | 1 | 3.18 | 15.5 | 15.5 |
| Inv. Temp. | 1 | 3.01 | 12.3 | 12.3 |
| Q-decay | 1 | 3.69 | 5.2 | 5.2 |
| Internalizing - Distress | 12 | 2.84 | 11.5 | 75.2 |
| SUIC__inq_grand_sum | 1 | 3 | 56.7 | 73.1 |
| **Externalizing - Antisocial/Disinhibition** | 10 | 4.05 | 14.3 | **46** |
| **Externalizing - Antagonism** | 8 | 5.06 | 5.3 | **27.7** |
| **SUIC__gcsq_fearlesness_of_death** | 1 | 6.37 | 7.2 | **24.5** |
| SUIC__gcsq_perceived_capability | 1 | 7.79 | 1.1 | 13.1 |
| Externalizing - Anankastia | 5 | 6.54 | 1.4 | 12.1 |
| DEMO__age | 1 | 8.11 | 1.5 | 9.9 |
| DEMO__sex_assigned | 1 | 7.86 | 0.4 | 8.7 |
| Internalizing - Fear | 13 | 6.54 | 0.3 | 6.3 |
| SUIC__gcsq_pain_tolerance | 1 | 9.83 | 0.2 | 2.3 |
| DEMO__active_mental_health_treatment | 1 | 10.02 | 0.1 | 1.1 |

*Table 19 Dataset II -Population level bootstrapping. Shown are the statistics across run for all computational, clinical and demographic categories. We computed the number of variables per category, the average rank across all runs, the percentage of runs in which the category scored highest among either computational or clinical/demographic category and the percentage of runs in which the category scored within the top 20 percent of categories based on the absolute weight on the first clinical covariate. Statistically significant category contributions are bolded for computational or clinical/demographic category separately.*

| Variable/Factor | N_variables | Mean_Rank | Rank1_Pct | Top20_Pct |
|---|---|---|---|---|
| **Escape Bias** | 1 | 2.05 | 49.7 | **49.7** |
| Avoid Bias | 1 | 3.06 | 17.3 | 17.3 |
| Learning Rate | 1 | 3.18 | 15.5 | 15.5 |
| Inv. Temp. | 1 | 3.01 | 12.3 | 12.3 |
| Q-decay | 1 | 3.69 | 5.2 | 5.2 |
| **Internalizing - Distress** | 12 | 2.84 | 11.5 | **75.2** |
| **SUIC__inq_grand_sum** | 1 | 3 | 56.7 | **73.1** |
| **Externalizing - Antisocial/Disinhibition** | 10 | 4.05 | 14.3 | **46** |
| **Externalizing - Antagonism** | 8 | 5.06 | 5.3 | **27.7** |
| **SUIC__gcsq_fearlesness_of_death** | 1 | 6.37 | 7.2 | **24.5** |
| SUIC__gcsq_perceived_capability | 1 | 7.79 | 1.1 | 13.1 |
| Externalizing - Anankastia | 5 | 6.54 | 1.4 | 12.1 |
| DEMO__age | 1 | 8.11 | 1.5 | 9.9 |
| DEMO__sex_assigned | 1 | 7.86 | 0.4 | 8.7 |
| Internalizing - Fear | 13 | 6.54 | 0.3 | 6.3 |
| SUIC__gcsq_pain_tolerance | 1 | 9.83 | 0.2 | 2.3 |
| DEMO__active_mental_health_treatment | 1 | 10.02 | 0.1 | 1.1 |

For cluster 1, bootstrapping suggests a more stable contribution of learning rate and avoid bias over escape bias (table 20), which had the highest weight in the full analysis. This lack of stability in the behavioural axis warrants caution in the clinical interpretation of the results and mirrors the lack of significance of the CCA for dataset I.

For dataset II, bootstrapping does confirm the clinical picture, where top contributing variables from the full analysis consistently score high in the Top 20% ranks among all clinical variables (cognitive problems: 71.1% , checking: 46.2%, non-persistence: 60.7, disorganization:45.9%, antisocial behaviour: 48.9%, anxious worry: 35.8%, NSSI: 38.2%, callousness: 25.4%), with the exception of hoarding (16.5%). At the subfactor level, this is reflected by significant loadings on internalizing distress and externalizing antisocial traits as well as the INQ score (Table 21).

*Table 20 Dataset I - Cluster 1 bootstrapping results. Shown are the statistics across run for all computational, clinical and demographic categories. We computed the number of variables per category, the average rank across all runs, the percentage of runs in which the category scored highest among either computational or clinical/demographic category and the percentage of runs in which the category scored within the top 20 percent of categories based on the absolute weight on the first clinical covariate. Statistically significant category contributions are bolded for computational or clinical/demographic category separately.*

| Subfactor | N_variables | Mean_Rank | Rank1_Pct | Top20_Pct |
|---|---|---|---|---|
| **Learning Rate** | 1 | 2.81 | 29.6 | **29.6** |
| **Avoid Bias** | 1 | 2.69 | 26.7 | **26.7** |
| **Inv. Temp.** | 1 | 2.73 | 23.9 | **23.9** |
| Escape Bias | 1 | 3.15 | 12.2 | 12.2 |
| Q-decay | 1 | 3.62 | 7.6 | 7.6 |
| **C_SSRS_ideation** | 1 | 5.65 | 31.5 | **47.8** |
| **BDI** | 1 | 4.89 | 15.6 | **47** |
| **BAI-T** | 1 | 5.29 | 9.1 | **38.9** |
| **GAD7** | 1 | 5.79 | 9.3 | **38.3** |
| **C_SSRS_ideation_sev** | 1 | 7.31 | 10.2 | **28.8** |
| STHS-state | 1 | 7.93 | 5.4 | 18.2 |
| STHS-trait | 1 | 8.06 | 4.7 | 15.2 |
| C_SSRS_attempt_no | 1 | 8.82 | 4.6 | 14.7 |
| C_SSRS_behav | 1 | 7.43 | 2.6 | 13.9 |
| BIS11 | 1 | 8.71 | 2.6 | 11 |
| C_SSRS_ideation_sev_pm | 1 | 8.38 | 1 | 8.5 |
| BPAQ | 1 | 8.72 | 1 | 7.1 |
| sex | 1 | 10.01 | 1.2 | 4.8 |
| ACE | 1 | 11.19 | 0.7 | 3.9 |
| age | 1 | 11.8 | 0.5 | 1.9 |

*Table 21 Dataset II - Cluster 1 bootstrapping results. Shown are the statistics across run for all computational, clinical and demographic categories. We computed the number of variables per category, the average rank across all runs, the percentage of runs in which the category scored highest among either computational or clinical/demographic category and the percentage of runs in which the category scored within the top 20 percent of categories based on the absolute weight on the first clinical covariate. Statistically significant category contributions are bolded for computational or clinical/demographic category separately.*

| Subfactor | N_variables | Mean_Rank | Rank1_Pct | Top20_Pct |
|---|---|---|---|---|
| **Learning Rate** | 1 | 2.81 | 29.6 | **29.6** |
| **Avoid Bias** | 1 | 2.69 | 26.7 | **26.7** |
| **Inv. Temp.** | 1 | 2.73 | 23.9 | **23.9** |
| Escape Bias | 1 | 3.15 | 12.2 | 12.2 |
| Q-decay | 1 | 3.62 | 7.6 | 7.6 |
| **Internalizing - Distress** | 12 | 2.96 | 16.8 | **70.2** |
| **Externalizing - Antisocial/Disinhibition** | 10 | 3.04 | 28.8 | **66.1** |
| **SUIC__inq_grand_sum** | 1 | 4.75 | 29.2 | **49.9** |
| **Externalizing - Antagonism** | 8 | 4.46 | 8 | **37.1** |
| **SUIC__gcsq_fearlesness_of_death** | 1 | 6.66 | 6.1 | **20.2** |
| Externalizing - Anankastia | 5 | 5.94 | 3.4 | 17.4 |
| Internalizing - Fear | 13 | 5.62 | 1.2 | 12.2 |
| DEMO__sex_assigned | 1 | 8.05 | 2.1 | 10.4 |
| DEMO__age | 1 | 8.46 | 3 | 8.1 |
| SUIC__gcsq_perceived_capability | 1 | 9.09 | 0.5 | 4.2 |
| DEMO__active_mental_health_treatment | 1 | 9.35 | 0.4 | 2.3 |
| SUIC__gcsq_pain_tolerance | 1 | 9.63 | 0.5 | 1.9 |

For cluster 2, bootstrapping confirms strong contributions from the Q-decay parameter on CV1 (fitted jointly across datasets, table 22). While the frequency of avoid bias and learning rate emerging as top contributors do not retain significance, they're rank order reflects the behavioural axis identified in the main analysis.

For dataset I, bootstrapping confirms strong contributions from depression and anxiety (table 22). Ideation severity is replaced by a significant contribution of suicidal ideation over bootstrap runs, whereas hopelessness does not retain significance. Overall, these results indicate the validity of the overall clinical correlations but warrants reproduction in larger sample sizes to understand the nuances of the observed patterns.

For dataset II, the GCSQ measures are most strongly indicated as top contributors, highlighting the role of constructs related to suicidal capability in this canonical variate. Additional significant contributions came from active mental health treatment (which was vanishingly small in the main analysis) and INQ. While the specific internalizing and externalizing contributions are spread across different factors and diverge from the main analysis, the pattern of correlation with suicidal capability persist through bootstrapping validation (table 23).

*Table 22 Dataset I - Cluster 2 bootstrapping results of 1000 bootstrap runs. Shown are the statistics across run for all computational, clinical and demographic categories. We computed the number of variables per category, the average rank across all runs, the percentage of runs in which the category scored highest among either computational or clinical/demographic category and the percentage of runs in which the category scored within the top 20 percent of categories based on the absolute weight on the first clinical covariate. Statistically significant category contributions are bolded for computational or clinical/demographic category separately.*

| Parameter | N_variables | Mean_Rank | Rank1_Pct | Top20_Pct |
|---|---|---|---|---|
| **Q-decay** | 1 | 2.28 | 43.5 | **43.5** |
| Avoid Bias | 1 | 2.92 | 19.7 | 19.7 |
| Learning Rate | 1 | 3.1 | 15.7 | 15.7 |
| Escape Bias | 1 | 3.28 | 14.5 | 14.5 |
| Inv. Temp. | 1 | 3.41 | 6.6 | 6.6 |
| **BDI** | 1 | 5.33 | 25.5 | **48.2** |
| **C_SSRS_ideation** | 1 | 6.09 | 19.2 | **38.5** |
| **BAI-T** | 1 | 7.44 | 7.3 | **23.6** |
| C_SSRS_behav | 1 | 7.81 | 7.8 | 21.9 |
| ACE | 1 | 8.1 | 5.2 | 20.7 |
| STHS-state | 1 | 7.79 | 6.1 | 19.9 |
| BIS11 | 1 | 8.05 | 4 | 18.3 |
| C_SSRS_ideation_sev | 1 | 8.16 | 4.1 | 18.3 |
| C_SSRS_ideation_sev_pm | 1 | 8.13 | 3.2 | 17.8 |
| sex | 1 | 8.54 | 6.7 | 15.9 |
| age | 1 | 8.57 | 2.1 | 13.8 |
| STHS-trait | 1 | 8.71 | 2.5 | 13 |
| GAD7 | 1 | 8.74 | 3 | 11.4 |
| C_SSRS_attempt_no | 1 | 8.96 | 1.6 | 11 |
| BPAQ | 1 | 9.55 | 1.7 | 7.7 |

*Table 23 Dataset II - Cluster 2 bootstrapping results. Shown are the statistics across run for all computational, clinical and demographic categories. We computed the number of variables per category, the average rank across all runs, the percentage of runs in which the category scored highest among either computational or clinical/demographic category and the percentage of runs in which the category scored within the top 20 percent of categories based on the absolute weight on the first clinical covariate. Statistically significant category contributions are bolded for computational or clinical/demographic category separately.*

| Parameter | N_variables | Mean_Rank | Rank1_Pct | Top20_Pct |
|---|---|---|---|---|
| Q-decay | 1 | 2.28 | 43.5 | **43.5** |
| Avoid Bias | 1 | 2.92 | 19.7 | 19.7 |
| Learning Rate | 1 | 3.1 | 15.7 | 15.7 |
| Escape Bias | 1 | 3.28 | 14.5 | 14.5 |
| Inv. Temp. | 1 | 3.41 | 6.6 | 6.6 |
| **SUIC__gcsq_pain_tolerance** | 1 | 4.42 | 28.4 | **57.4** |
| **SUIC__gcsq_fearlesness_of_death** | 1 | 4.98 | 24.2 | **49.6** |
| **SUIC__gcsq_perceived_capability** | 1 | 5.54 | 14.2 | **42.5** |
| **DEMO__active_mental_health_treatment** | 1 | 6.35 | 8.8 | **33** |
| **SUIC__inq_grand_sum** | 1 | 6.78 | 11.1 | **28.7** |
| **Externalizing - Anankastia** | 5 | 5.63 | 2.7 | **25** |
| DEMO__sex_assigned | 1 | 8.68 | 2.7 | 13.2 |
| Externalizing - Antisocial/Disinhibition | 10 | 6.87 | 0.7 | 12.7 |
| DEMO__age | 1 | 8.31 | 3.1 | 12.6 |
| Internalizing - Distress | 12 | 7.02 | 1.8 | 8.7 |
| Internalizing - Fear | 13 | 6.56 | 0.6 | 8.6 |
| Externalizing - Antagonism | 8 | 6.88 | 1.7 | 8 |
